# Identifying Optimal Parameters for Neuroscience-Informed Interventions for Misophonia

**DOI:** 10.1101/2023.06.25.23291872

**Authors:** Andrada D. Neacsiu, Lysianne Beynel, Nimesha Gerlus, Victoria Szymkiewicz, Kevin S. LaBar, Noreen Bukhari-Parlakturk, M. Zachary Rosenthal

**Author notes:** Correspondence concerning this article should be addressed to Andrada D. Neacsiu, Duke University Medical Center (102505), Durham, NC, 27710. **Fields explored:** psychotherapy and psychopathology (primary), general topics in psychiatry (secondary).

## Abstract

**Background:** Misophonia is the inability to tolerate certain aversive, repetitive common sounds.

**Methods:** Using a within-subjects experimental design, twenty-nine participants with misophonia and thirty clinical controls with high emotion dysregulation received inhibitory neurostimulation (1Hz) over a personalized medial prefrontal cortex (mPFC) target functionally connected to the left insula; excitatory neurostimulation (10Hz) over a personalized dorsolateral PFC (dlPFC) target; and sham stimulation over either target. Stimulations were applied while participants were either listening or cognitively downregulating emotions associated with personalized aversive, misophonic, or neutral sounds. Subjective units of distress (SUDS) and psychophysiological measurements (skin conductance response[SCR] and level [SCL], and high-frequency heart rate variability [HF-HRV]) were collected.

**Results:** Compared to controls, participants with misophonia reported higher distress (Δ_SUDS_ = 1.91-1.93, *p*s<.001) when listening to and when downregulating misophonic distress, although no psychophysiological differences were found. Both types of neurostimulation reduced distress significantly more than sham, with excitatory rTMS providing the most benefit (Cohen’s *d_SUDS_=* 0.53; *d*_SCL_= 0.14). Excitatory rTMS also enhanced the regulation of emotions associated with misophonic sounds in both groups when measured by SUDS (*d_control_*= 1.28; *d_Misophonia_=* 0.94), and in the misophonia group alone when measured with SCL (*d*= 0.20). Both types of neurostimulation were well tolerated and feasible to administer.

**Discussion:** Clinical controls and misophonic participants were different in their self-report but not in psychophysiological measures of distress and regulations. Participants reported the lowest misophonic distress when engaging in cognitive restructuring enhanced with high-frequency neurostimulation, a finding that offers insight into the best path forward for misophonia interventions.

## Introduction

Misophonia is an intolerance for certain aversive repetitive and common sounds such as chewing, swallowing, or keyboard tapping. When presented with those sounds, individuals with misophonia experience intense distress associated with heightened physiological reactions such as increased heart rate or skin conductance and have trouble disengaging from the trigger [1]. While the prevalence of misophonia is estimated at 5-12% of the population [2], this condition is still not well understood, and the first consensus on a standardized definition was published only recently [3]. Current misophonia treatment approaches include lifestyle modification, cognitive behavioral therapy, and audiological treatment. However, since the mechanisms underlying misophonia are unknown, the current treatments do not have clear targets for change; limited evidence exits on the benefits of any interventions; and misophonic patients report limited satisfaction with many common approaches [4].

Emerging neuroscientific findings suggest that misophonia may be an independent disorder [5], characterized by hyperactivity of the anterior insular cortex (AIC) in response to misophonic trigger stimuli [1,6], and altered activity and connectivity of prefrontal structures. These findings suggest enhanced salience of and difficulty disengaging from misophonic sounds. Qualitative studies report that the misophonic experience contains intense emotions of anxiety, anger, and disgust, among other emotions [7–10], as well as difficulties with emotional regulation [11,12]. Thus, examinations of emotional dysregulation and comparisons with clinical populations who have difficulty with their arousal and regulation are warranted. Although several candidate networks are at play in this complex disorder (e.g., sensory, auditory, or even motor [13]), examining the similarities and differences of misophonia with other emotional disorders holds promise because of the wealth of existing interventions for emotional dysregulation across psychopathology [14–16].

In the present study, we compared transdiagnostic clinical adults who report high emotional dysregulation with adults with moderate to severe misophonia using multimethod assessment. Specifically, we examined self-report and psychophysiological differences when responding to neutral, non-misophonic aversive, and personalized misophonic trigger sounds. We also examined differences between groups when engaging in the downregulation of misophonic or aversive sounds using cognitive restructuring [17,18]. We examined differences in the response and regulation of aversive sounds to test broad emotional dysregulation in misophonia. Furthermore, we tested how clinical controls respond to and regulate emotions associated with trigger sounds to clarify the unique features of misophonia.

In addition, we aimed to investigate whether a bottom-up versus a top-down approach is best suited for a misophonia intervention. Using repetitive transcranial stimulation (rTMS) [19], we examined whether inhibiting insula activity with low-frequency mPFC stimulation or enhancing regulation with high-frequency dlPFC stimulation during the presentation of misophonic sounds leads to larger reductions in misophonic distress when compared to listening to trigger sounds without any intervention. We also wanted to compare the use of cognitive restructuring (an emotional regulation skill), with neurostimulation, and with skill use plus neurostimulation as different avenues for treatment development using a within-subject design. Our goal was to answer several questions to accelerate the development of treatments for misophonia.

RTMS is an evidence-based non-invasive technique that employs transient magnetic fields to induce an electrical current in the cortex and to temporarily alter neuronal activation or connectivity in a targeted circuit [20–22]. Over time, rTMS is thought to have a therapeutic effect by inducing lasting changes in synaptic plasticity in targeted networks [19]. Optimizing neurostimulation protocols, such as target selection, is known to improve intervention efficacy. Stimulation guided by functional neuroimaging is more effective than traditional targeting [23,24], with almost doubled effect sizes [25]. Further, to enhance emotion regulation with neurostimulation, findings point to the right dlPFC as a promising target [26]. Other findings also suggest that reducing intra-train intervals for excitatory neurostimulation may improve outcomes [27].

Targeting deeper brain structures, such as the insula, requires connectivity-based rTMS because the electric field directly penetrates only up to approximately 3 cm into the cortex [28]. A growing number of studies have demonstrated that neurostimulation reaches distal brain regions via their connections with superficial cortical regions directly targeted. There has been a steady growth in the number of studies demonstrating that “connectivity-based rTMS” can be used to successfully modulate the activity of deep brain regions such as the amygdala [22,29,30], the insula [31], or the anterior cingulate cortex [32]. Given prior findings of hyperconnectivity between the insula and the medial prefrontal cortex (mPFC) in misophonia [1], the mPFC was selected as the cortical target over which low-frequency rTMS (LF-rTMS) was applied to inhibit activity in the left insula.

We trained all participants to use cognitive restructuring (CR) to understand whether skills training could be as effective for misophonic adults as it is for clinical emotional dysregulation [15]. CR is a fundamental skill in many evidence-based psychotherapies [33–35] that also has been studied in basic neuroscientific research [18,36]. CR involves thinking differently about a situation to change the emotional experience and is currently one of the most studied emotional regulation strategies across both basic and clinical research [37]. Functional neuroimaging studies examining CR have found activation distributed throughout a fronto-limbic network, including the dlPFC, mPFC, amygdala, and insula [18,36,38], providing support for the use of this intervention in misophonia.

Effective use of reappraisal during an emotion regulation task increases high-frequency heart rate variability (HF-HRV) [39], a marker of effective emotion regulation [40,41]. HF-HRV has not yet been investigated in misophonia. Rather, changes in skin conductance level (SCL) and response (SCR) were identified as potential psychophysiological markers of misophonic distress [1]. There have been few examinations of the effect of emotion regulation tasks on SCL, with one study showing no differences in SCL between effective and maladaptive regulation strategies [42].

Therefore, one aim of this study was to test differences between adults with misophonia and clinical controls on HF-HRV, SCL, SCR, and self-reported distress (SUDS) during passive listening and regulation of aversive and misophonic sounds. The second aim was to examine whether potential misophonia interventions should target reduction to sound reactivity or improvement in emotion regulation as the primary mechanism of change. Furthermore, we examined whether neurostimulation, cognitive restructuring, or their combination offers the most promise for a novel intervention.

We hypothesized that when compared to controls, participants with misophonia will exhibit (H1) higher distress (as measured with SUDS, SCL, and SCR) when listening to misophonic trigger sounds; (H2) lower distress when listening to aversive sounds; and (H3) lower HF-HRV during the regulation of misophonic sounds. We expected that both LF-rTMS (H4) and HF-rTMS (H5) stimulation will reduce distress significantly more than sham stimulation during the presentation of misophonic cues in the misophonia group. We expected HF-rTMS stimulation to lead to lower distress and higher regulation (HF-HRV) in both groups when compared to sham stimulation during the presentation and regulation of aversive sounds (H6). We planned to explore differences between the use of CR, LF-rTMS, HF-rTMS, and their combination (i.e., CR+HF-rTMS; CR+LF-rTMS) within and across groups, expecting combined interventions to lead to more reductions in arousal that each component alone (H7).

## Methods

### 1. Participants and Procedures

This study was pre-registered under the Clinical Trials ID (NCT04348591) and ran between October 2020 and May 2022. The study was powered based on expected effect sizes derived from Kumar et al. [2], who compared adults with misophonia (*N* = 20) with healthy controls (*N* = 22) and found significant differences in AIC response, with an effect size of 1.27 (mean difference between groups = 2.7 a.u, *SD* = 2.11). For the proposed design, with planned comparisons across condition (downregulate vs. listen), sound type (misophonic, aversive, or neutral), and group (misophonic vs. clinical control), an expected effect size of 1.27, and alpha error rate set at .05, we expected adequate power (over 80%) with 29 participants in each group. Therefore, we aimed to recruit 30 participants in each condition.

Participants were recruited through online websites (e.g., Craigslist, Dukelist, CMER), social media (Facebook, Reddit), researchMatch, flyers, and electronic medical record outreach. Interested participants completed an online and in-person screen before being invited for an imaging session. The CONSORT diagram (Figure 1) and the Supplement contain additional details regarding enrollment in the study.

**Figure 1.**
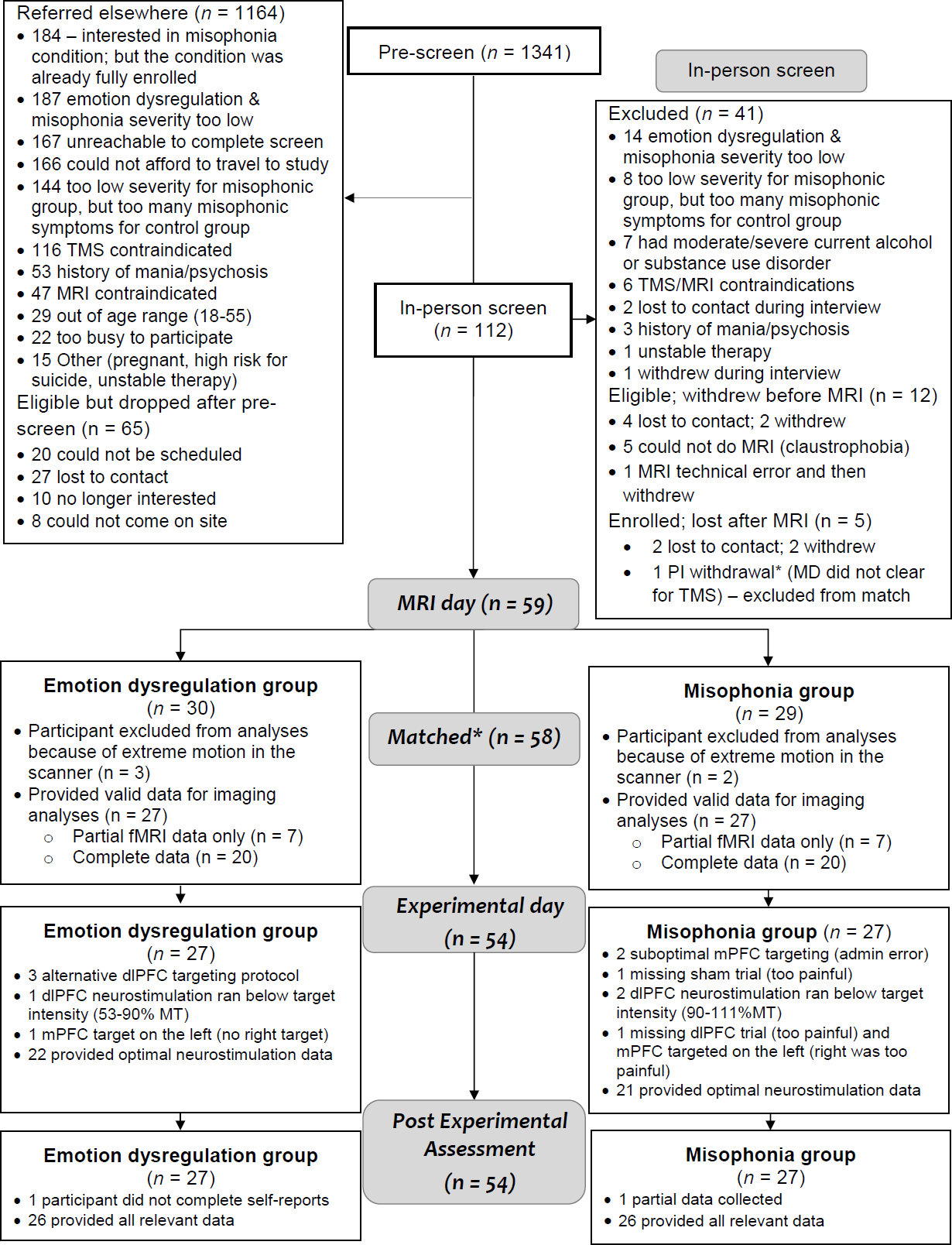
CONSORT Diagram depicting study flow

Enrolled participants were 4 men and 52 women and 3 non-binary adults between the ages of 18 and 54 (*M* = 28.31; *SD* = 9.06 years old) who self-reported significant misophonic severity, or who met criteria for any DSM-5 disorder (excluding active substance use, psychotic disorders, and Bipolar I) and self-reported above average emotion dysregulation. Participants met criteria for an average of 1.75 (*SD =* 1.62) current diagnoses and 3.54 (*SD* = 2.26) lifetime diagnoses according to the structured interview for *DSM-5* disorders (SCID-5) [43] (see Table 1). Twenty-one participants (36.21%) met criteria for at least one personality disorder according to the SCID-5-PD [44].

Significant misophonic severity was operationally defined as a score of 2 or higher on both the sensitivity to trigger sounds and behavioral reactivity subscales of the misophonia questionnaire (MQ)[2] and a score of 7 or higher on the MQ severity scale. High emotion dysregulation was operationally defined as a total score of 89 or higher on the Difficulties in Emotion Regulation Scale (DERS) [45] as established in other prior studies [25]. Misophonic participants were not excluded if they reported heightened emotion dysregulation, but control participants were excluded if they reported significant misophonic symptoms on the MQ. Additional exclusion criteria are presented in Figure 1. Participants received a maximum compensation of $250. The study was approved by the Duke University Health System Institutional Review Board. Figure 2 depicts the study design.

**Figure 2.**
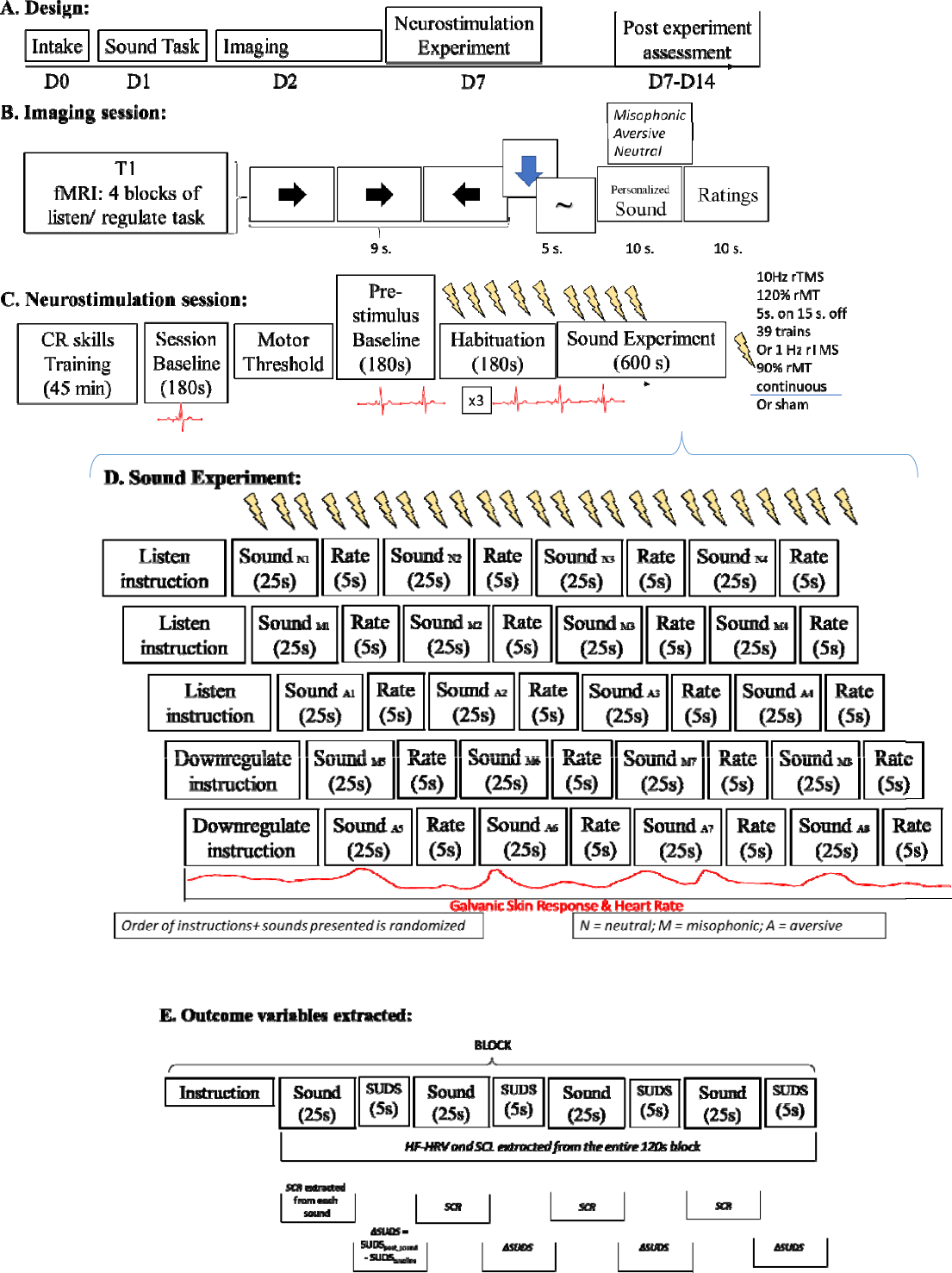
Experimental design. D represents the day in the study when these sessions were approximately scheduled. CR = Cognitive Restructuring; rTMS = repetitive transcranial magnetic stimulation; MT = motor threshold

### 2. Intake Session

After providing voluntary, written informed consent, participants completed diagnostic assessments (SCID-5, SCID-PD), a verbal intelligence test [46], and a questionnaire packet.

### 3. Measures

The total score from the *<u>Difficulties in Emotion Regulation Scale</u>* (DERS) was employed to measure the severity of emotion dysregulation. The *<u>Misophonia Questionnaire (MQ)</u>* assessed the severity of misophonic distress. The feasibility and acceptability of the experimental visits were evaluated using two questionnaires administered before and after each experimental task (sounds, MRI, neurostimulation). Specifically, we asked participants to rate their current level of distress (rated on a scale from 1-7 [50]) and their state negative affect (*<u>Positive and Negative Affect Scale;</u>* PANAS [48]). Further details on these scales (Cronbach’s alpha, inter-rater reliability) can be found in the Supplement.

During each visit, participants were asked several times to rate dissociation using a 4-item scale [49] as well as their current distress on the *<u>Subjective Units of Distress Scale</u>* (SUDS; range: 0 [no distress] to 9 [extreme distress] [47]). Before and after the neurostimulation experiment (and after each type of neurostimulation was administered), participants rated on a scale from 0–3 (absent, mild, moderate, severe) the intensity of their headache, neck pain, scalp pain, seizure (as observed by the technician), hearing impairment and any other side effect that they might have experienced from the TMS treatment. The success of the blind was measured using a scale from 1 (I am certain I received sham stimulation) to 9 (I am certain I received active stimulation). A rating of 5 indicated uncertainty.

Finally, an unpublished, previously developed interview [51] was used to examine feasibility and acceptability as directly relevant to the study. See the supplementary file for question topics and scoring.

### 4. Sound Task

After intake, qualifying participants completed an in-person baseline task where they were seated in front of a computer in a sound-attenuated room with noise-canceling headphones on. All participants heard 101 pre-selected sounds: 31 aversive, 40 misophonic, and 30 neutral sounds. Sounds were selected from the International Affective Digitized Sound System (IADS [20]; n = 64 sounds), from a freely available sound database (https://www.fesliyanstudios.com/royalty-free-sound-effects-download; n = 12 sounds), as well as from published literature (n = 25 sounds) [1]. The IADS is a standardized set of *\*.wav* files that have been normed for arousal and valence levels. Non-misophonic aversive and neutral stimuli for this task were selected using the standardized values of valence and arousal from the IADS. Misophonic sounds were selected using the literature, to include sounds that have been previously described as being particularly triggering for misophonic individuals. To make sounds from various sources consistent, each sound was cut or repeated for a total play duration of 6 s. Each stimulus presentation was followed by valence (positive, negative, or neutral) and arousal ratings, and then a 6-s return-to-baseline period. Participants in the misophonia group alone were additionally asked whether the aversive sound was a misophonic trigger for them (yes/no).

Based on this task, for each participant, personalized sets of 12 aversive, 12 misophonic, and 12 neutral sounds were selected for the neurostimulation and neuroimaging sessions. For the misophonia group, the highest negative arousal sounds that were flagged as misophonic triggers were included in the misophonic sounds condition. The highest negative arousal sounds that were not marked as misophonic triggers were included in the aversive condition. For the control group, the highest negative arousal sounds that were initially labeled as potentially misophonia related created the misophonic sounds, the highest negative arousal sounds that were from either the original aversive or neutral groups were selected for the aversive condition.

These sounds were shuffled so they would play in different orders during the imaging and the neurostimulation sessions. Sounds were repeated during the neurostimulation session, once for each run (i.e., the same sounds were played during different neurostimulation conditions, but in a different order) to examine the intervention potential of each type of neurostimulation on similar stimuli. The pseudo-randomized order ensured that the same sound would not play back-to-back in two different runs. See supplement for additional details.

### 5. Neuroimaging sessions

#### MRI training

Participants completed an imaging session (Figure 2B) on average 9.83 days (SD = 9.97) after the eligibility intake session. During the first hour of this session, participants were introduced to the task stimuli and were trained in how to respond accordingly: simply look at the screen when a cross was presented (passive baseline); indicate with a button press the direction of an arrow shown on the screen (active baseline); prepare to downregulate emotions associated with the sounds they hear when seeing a downward arrow; and prepare to fully engage with listening and allowing any experience that arises during a sound file when seeing a wave symbol on the screen (∼).

Participants were then introduced to CR as a strategy to use when instructed to downregulate emotions. Specifically, participants were told, *“When you see the downward arrow, prepare to reduce negative emotions when you hear the following sound”.* Restructuring techniques including distancing and reframing tactics were briefly introduced and practiced with two standardized aversive sounds. Participants were told to reinterpret the context of the sound in some way (e.g., instead of focusing on a baby’s cry by thinking that the baby is distressed, think that someone will come to pick the baby up; reframing technique). Alternatively, participants could think that the sound is part of a movie and not a real sound (distancing technique). Participants were free to pick either tactic when prompted to downregulate. These tactics were briefly introduced during a 10-minute discussion to ensure the participant understood how to engage in the targeted behavior during the scan, but not such that the participant learned new ways to downregulate emotions.

#### MRI acquisition

After training, participants moved into a research-dedicated GE HD 3.0 T MRI scanner. The scan started with an anatomical image (3D MPRAGE pulse sequence; time repetition [TR] = 2300 ms; echo time [TE] = 3.2 ms; image matrix = 2562; flip angle = 12°; voxel size = 1-mm isotropic; 162 contiguous axial slices). Field maps were collected for half of the sample (depending on the time available during the scan) to correct for the offline distortion of BOLD images. Four runs of EPI functional images were then acquired utilizing in-plane and multi-band acceleration, allowing the collection of high-resolution functional data while minimizing spatial distortion and signal dropout related to magnetic susceptibility (SENSE acceleration factor = 1; multi-band factor = 3; TR = 2000 ms; TE = 30 ms; image matrix = 1282; flip angle = 77°; voxel size = 2-mm isotropic; 68 contiguous axial slices).

Functional runs were acquired while participants experienced and downregulated emotions related to personalized sounds (task-related acquisition: four runs). Each run included five blocks (3 trials per block) presented in a pseudo-random order (downregulate aversive, downregulate misophonic, hear neutral, hear misophonic, hear aversive). In each trial, participants first observed a fixation cross (3-7s) followed by the active baseline task. Then, participants were cued to use a strategy (downregulate or hear). Next, they were presented with a misophonic, aversive, or neutral sound stimulus. Participants rated their subjective distress after each trial. Participants wore earphones to hear the sounds and to attenuate scanner noise. Head motion and further outside sound exposure were minimized with foam pads placed on both sides of the participant’s head. Data from this imaging session were used to develop the neurostimulation targets.

#### Randomization

Following the MRI visit, the sequence of targets to be used during the neurostimulation session was randomized using an Excel number generator for participants who still qualified. Each participant received active neurostimulation over the right dlPFC, active over the right mPFC, and sham over either the right dlPFC or mPFC (randomly determined). The subject and the PI were blinded to the neurostimulation condition (active versus sham) for all trials. The technician who assisted with the session and prepared the equipment was not blind to the target order and was responsible for following the randomization protocol during the neurostimulation visit. The technician did not influence in any way the course of the neurostimulation experiment and minimally interacted with the participant.

#### Neuroimaging Data Analysis

FMRI analyses were performed immediately after the MRI session to define the individualized stimulation target. Structural and functional data were first examined with MRIQC for quality assurance and then preprocessed with fMRIprep v1.1.4 [52]. The non-aggressive variant of ICA-based automatic removal of motion artifacts (AROMA) [53] was also applied to the fMRI data. After this preprocessing by fMRIprep, the fMRI data underwent high-pass temporal filtering with a 100-second cutoff (FSL’s *fslmaths*) and were masked with a standard MNI brain template to exclude erroneous signals outside the brain. At the first level, functional data were analyzed as individual runs, using a general linear model (GLM) implemented in FSL’s FEAT procedure [55]. Model regressors were created for each task block, instruction cue, and active baseline task (see supplemental file).

The “downregulate vs. listen to a misophonic sound” contrast used for dlPFC targeting was defined as part of a participant-level fixed effects model. A psychophysiological interaction (PPI) analysis was also conducted to measure functional connectivity between the left insula and the right mPFC during the “listen to misophonic sound vs. neutral sound.” The left insula was defined by the anatomical mask available as part of the FSL Harvard-Oxford sub-cortical structural segmentations. Statistical PPI maps from each of the four runs were then combined into a second-level analysis using a fixed-effect model for each person.

#### Target Identification

The statistical maps for the contrasts of interest (“downregulate vs. listen to a misophonic sound”, and “PPI: listen to misophonic vs. neutral sound”) were transferred to native space back from standard space using the inverse ANTS transformation [56], and overlaid onto the anatomical image in neuronavigation software (BrainSight, Rogue Research, Canada). For each participant, the cluster within the right dlPFC showing the strongest positive z-statistic value for the “Downregulate” contrast was defined as the dlPFC target (average z_dlPFC_stimulation_site_ = 2.74, SD = .73). The cluster within the right mPFC showing the strongest positive z-value above a threshold of z = 1.96 for the PPI-“Listen” contrast was defined as the AIC functional connectivity target(average z_mPFC_stimulation_site_ = 3.03, SD = 1.50). Examples of how targets were extracted for five individuals in each group are included in Figure 3A.

**Figure 3A.**
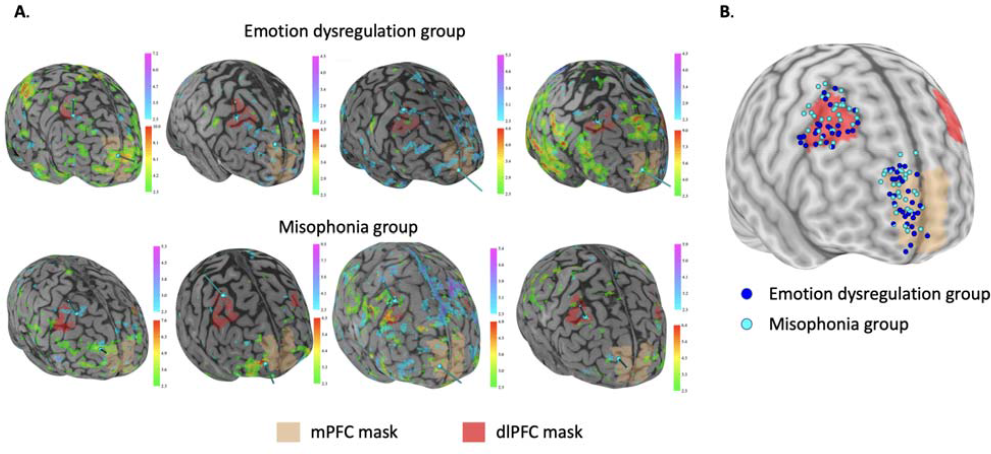
The top legend is the range for activation for the regulation contrast; regulation contrast is depicted with a blue-magenta map; the bottom legend is the range of activation for the reactivity contrast, depicted with the green-red map. The copper region is the structural mPFC mask used to constrain targets. The red region is the dlPFC mask used to constrain the regulation targets. All images are in native space and are thresholded by a minimum z of 2.3. **1B**: Coil position across all participants, with in dark blue the emotion dysregulation group, and light blue the misophonia group.

The coil was manually oriented so that the induced E-field was perpendicular to the closest sulcal wall. When this orientation was not feasible (e.g., coil handle in front of participant’s face), the orientation was changed by 180 degrees, and the current was reversed. Scalp-to-cortex distance in mm at each site of stimulation was measured using the tape tool in BrainSight. Hair thickness was measured using a digital depth gauge (Audew, HK, resolution: 0.01 mm) over the target (89). The brain-to-coil distance was computed as the sum of the hair thickness and the distance from the scalp to each target and was included as a covariate in the analyses given previous results that it may impact outcomes [25]. See **Figure 3B** for a visual depiction of all personalized targets.

### 6. Neurostimulation Experimental Session

Participants returned for the 3.5-hour skills training/neurostimulation session (Figure 2C) on average 14.11 days after the MRI session (SD = 12.57).

#### Skills training

The first 45 minutes were spent on skills training, one-on-one with the first author, a clinical psychologist with expertise in cognitive behavioral therapy. The session focused on in-depth learning of CR and on practicing it on standardized and personal examples. Skills training used standardized procedures blending psychotherapeutic approaches [57,58] with instructions in CR that matched prior neuroimaging studies [59]. Participants were told that thoughts and emotions are interconnected and one validated way to change emotional experiences is to reduce problematic thoughts in situations that prompt the emotions [58,60-62]. Participants were taught *distancing* and *reframing* tactics. Distancing by using objectivity, time, and space was discussed and practiced on standardized examples [10]. For the misophonia group, distancing was further refined as thinking about what else could be making that sound, thinking that the sound is time-limited, or trying to focus one’s attention in the presence of a trigger on another element of the situation that is not as upsetting. Participants were also taught *reframing* using an adapted version of existing paradigms [11–13]. For the misophonia group, we worked on reframing thoughts related to the person producing the sound (“they’re hungry rather than they’re rude”; “they’re anxious rather than they’re annoying me on purpose”). We also discussed using the three questions (worst case scenario, likelihood it worst case will happen, and likelihood of survival) during an experience where misophonic triggers are present. See more details in the Supplement.

#### TMS experiment

Following the successful completion of the behavioral training, the resting motor threshold (rMT) was established. Psychophysiological measurements were collected continuously during the experiment using the BIOPAC MP150 recording system (Goleta, CA) via GSR electrodes on the index and ring fingers as well as HR electrodes on the ankle and wrist.

Active and sham rTMS were performed with a figure-8 coil (A/P Cool-B65) and a MagPro X100 stimulator (MagVenture, Denmark) set up to deliver biphasic pulses. Ten Hz rTMS over the personalized right dlPFC target (HF-rTMS) was performed using 5 s of stimulation and 15 s of an inter-train interval (ITI) at 120% rMT. A frequency of 10 Hz has been successfully used in affective disorders [63–66], and an ITI of 15 s was chosen based on evidence suggesting the superiority of 15 s over 26 s [67].

One Hz rTMS over the personalized mPFC-AIC-connectivity target (LF-rTMS) was performed using one pulse per second continuously at 90% rMT [68]. Sham stimulation was applied using the same intensity setting but with the coil in placebo mode, which produced similar clicking sounds and somatosensory sensations (via electrical stimulation with scalp electrodes) without a significant magnetic field reaching the brain [69]. This type of sham stimulation allowed participants and the experimenter to stay blind to the type of stimulation they received.

Coil position and orientation were continually monitored through a stereotaxic neuro-navigation system (Brainsight, Rogue Research, Canada). All participants received the three interventions. Each neurostimulation experimental session was conducted by the first author (AN), with the assistance of a TMS technician. AN, who was blinded to the stimulation condition, led the participant through the session and decided on dose adjustments and course of action for any protocol deviations. For example, if a participant could not tolerate rTMS, the first author could decide to drop the intensity to rMT and slowly increase it during habituation.

The experimental session (Figure 2C) included a 180 s pre-session baseline, followed by three experimental runs, one using HF-rTMS over the right dlPFC (39 trains), one using continuous LF-rTMS over the right mPFC, and one using sham rTMS applied over one of those two targets, in a randomized order. Each run included a pre-task baseline (180s), a habituation period (180s) where neurostimulation was administered while the participant was not given any instruction, a sound task comprised of 5 blocks (120s each), and a 600 s break. Neurostimulation was administered throughout this sound task: each block started with an instruction (downregulate or listen) and was followed by four sounds each (either misophonic, neutral, or aversive depending on the block) played on repeat for 25 seconds. Sound and block order was randomized within each run. During the sound presentation, a downward arrow (for downregulate), or wave symbol (∼, for listen) was present on the screen to cue participants on whether to downregulate or listen (Fig. 2D).

#### Psychophysiological Data

High-frequency heart rate variability (HF-HRV), skin conductance response (SCR), and skin conductance level (SCL) were used as indicators, respectively, of emotion regulation, peak emotional arousal, and average arousal. AcqKnowledge software was used to collect and pre-process raw ECG data [70]. For each baseline, habituation period, and sound task block, high-frequency HRV (HF-HRV) was extracted from the ECG signal. HF-HRV requires at least 120s to be accurately assessed, precluding its measurement at each sound presentation; for this reason, the average value within each sound task block was employed as the outcome measure. Galvanic skin response (GSR) was recorded continuously in tandem with ECG. For each baseline, habituation period, and for each sound presentation we extracted the maximum GSR value (SCR). Using a MATLAB program, we derived the SCL from the GSR signal for each baseline, habituation period, and sound task block by leveling the signal when clear spikes were identified [71] to remove SCRs. Therefore, SCL was also extracted from 120s blocks, like HF-HRV (see Figure 2E for a visual depiction of variables extracted for analyses).

### 7. Statistical Analyses

To compare the effects of our neurostimulation conditions on emotion regulation, we conducted four analyses examining ΔSUDS, SCR, SCL, and HF-HRV (see Figure 2E). We planned to control for the values of these outcome variables during baseline. Before main analyses were conducted, we examined differences in main outcomes during habituation, where neurostimulation was administered in the absence of any other stimuli. If we found differences in outcomes during habituation, we planned to replace the baseline value with the outcome value during habituation. If no differences were found, the baseline covariate represented the outcome value during the task baseline.

For all statistical models, the experimental group (misophonic or emotion dysregulation) was included as a between-subject variable. Within-subject fixed effects included the experimental neurostimulation condition (active HF-rTMS over the right dlPFC, active LF-rTMS over the mPFC-insula, sham rTMS), the block instruction (downregulate misophonic, downregulate aversive, hear misophonic, hear aversive, hear neutral), the time within the experiment (0-2, corresponding with the runs employed), and the time within each run (0-4, corresponding to each block). The analysis also included the group by condition by instruction interaction effect. For SCR and SUDS analyses, the time within the block (0-3, corresponding to each of the four sounds played) was also examined as a fixed effect. Time was considered a categorical variable. To account for multiple comparisons, we used a Bonferroni correction and reduced the alpha threshold to less than .0125. Differences between LF and HF rTMS were not hypothesized but explored when both stimulation conditions were significantly different compared to sham.

Planned covariates for all analyses included coil-to-cortex distance given evidence that higher distance may impede the effectiveness of neurostimulation interventions for emotion regulation that are not designed to account for this parameter [72,73]. Data-driven covariates such as the incidence of headaches post-stimulation were also examined and added as needed. Effect sizes were computed by using Feingold’s formula [74] and interpreted using Cohen’s specifications [75].

#### Normality Assumption

All variables were tested to examine whether they violated the normality of distribution assumption using Shapiro-Wilk normality tests (*W* < .90). The distribution of HF-HRV was transformed to normal using the function lg10 (HF-HRV*1000000). All other data were normally distributed.

## Results

### 1. Feasibility, acceptability, tolerability, compliance, and blinding efficacy

#### Acceptability, feasibility, tolerability, and compliance

Overall participants in both groups reported reduced stress at the end compared to the beginning of each experimental session (Δ_average_intake_change_ = - 0.56, SD = 1.53; Δ_average_MRI_change_ = −0.31, SD = 1.22; Δ_average_TMS_change_= −0.48, SD = 0.89). These results suggest that the experimental tasks were feasible and acceptable to our participants. Headache was the most commonly reported side effect of the stimulation with no between-group differences (p = 1.00) in headache incidence. Following the sham stimulation, 26 participants (48.15%) reported some headache, compared to 40 participants (74.07%) after the HF-rTMS trial, and 34 (62.96%) after the LF-rTMS trial, a significant difference (χ [2] = 7.73, *p* = .02). Because of the higher incidence of headache in the active stimulation conditions, we included this variable as a data-driven covariate in analyses examining differences during the neurostimulation experiment. Neck pain and scalp discomfort were other side effects reported by participants. See supplementary materials for additional findings related to feasibility, tolerability, and compliance. Participants reported staying present during the experimental tasks (*M* = 7.18, *SD* = 1.75, range 0-9), and being successful in following the instructions (*M* = 7.50, *SD* = 1.60, range: 0-9) with no differences between groups, or between the type of neurostimulation received (active HF-rTMS, active LF-rTMS, or sham), ps > .17.

#### Blinding efficacy

Participants tended to believe they received active stimulation across runs (*M* = 6.30, *SD* = 2.21, Range: 0-I received sham; 5-unsure; 9 – I received active rTMS). There was no difference in guessing active versus sham assignments between types of neurostimulation received (*M_sham_*= 5.72, *SD* = 2.27; *M_HF-rTMS_*= 6.74, *SD* = 2.07; *M_LF-rTMS_* = 6.43, *SD* = 2.18) or between groups (F [2, 151] = 11.05, *p* = .09), according to a multivariate ANOVA controlling for racial background (F [1, 151] = 0.06, *p* = .44).

### 2. Preliminary Analyses Examining Confounds

There were no significant differences between groups in age (*t*[57] = −1.07; *p* = .30) or gender distribution (_χ_ [4] = 2.98, *p* = .56), suggesting that the matching was successful. There were also no differences between groups in sexual orientation, marital status, education, income, use of psychotropic medication, current enrolment in psychotherapy, presence of personality disorders, or the number of current or lifetime diagnoses met (*p*s > .05). There were significantly more participants belonging to racial or ethnic minority groups in the emotional dysregulation condition. Therefore, racial/ethnic background was recoded as 0 (non-white) or 1 (white) and co-varied in subsequent analyses). During the rTMS experiment, there was no significant difference in dissociation between groups or between types of neurostimulation (*F_group_*[1, 38.51] = 2.30, *p =* .14; *F_experimental_condition_* [2, 65.88] = 0.21, *p =* .81). There was also no significant difference at the beginning of the session in sleepiness between groups (*t*[52] = 0.52, *p* = .61). Therefore, these variables were not included as covariates and final MMANOVA models used task baseline value, racial background, presence of headache, and coil-to-cortex distance as covariates.

#### Psychophysiological effects induced by rTMS alone, during the habituation period

Significant differences were found between experimental conditions (sham vs. active rTMS) during the habituation period (i.e., when neurostimulation alone was administered at rest). At rest, HF-rTMS administered over the right dlPFC enhanced HF-HRV (*F* [2, 90.49] = 6.63, *p* = .002), SCR (*F* [2, 90.76] = 14.27, *p* < .001), and SCL (*F* [2, 96.22] = 27.17, *p* < .001) significantly more than sham neurostimulation (Δ_HF-HRV_ = 0.12, SE = .03; Δ_SCR_ = 2.84, SE = .54; Δ_SCL_ = 1.76, SE = .24). LF-rTMS over the right mPFC did not significantly affect HF-HRV, but significantly increased SCR (Δ_SCR_ = 1.72, SE = .52) and SCL (Δ_SCL_ = 0.72, SE = .23) when compared to sham. Participants experienced significantly higher tonic arousal (SCL) during HF-rTMS than during LF-rTMS (Δ_SCL_ = 1.05; SE = .24, *p* < .001).

Caucasian participants had a significantly lower HF-HRV across habituation periods when compared to non-Caucasian adults (*F_HF-HRV_*[1, 38.89] = 7.70, *p* = .008, Δ = .13, SE = .05). No other covariates (e.g., headache, coil-to-cortex distance) were significant across analyses. Taken together, these findings highlight that neurostimulation induces increased participants’ arousal across several psychophysiological measurements, beyond the discomfort captured by covariates. They also highlight the unique effect of excitatory neurostimulation over the right dlPFC in enhancing emotion regulation as assessed by heart rate variability. As planned, because of these observed differences, habituation values for outcomes variables replaced task baseline values as co-variates in main analyses. The SD for HF-HRV (0.59), SCR (4.26), and SCL (3.60) during the session baseline, and for SUDS post-session baseline (1.09) were used to compute the effect sizes.

### 3. The Effect of Neurostimulation and Cognitive Restructuring on Reactivity and Regulation

Table 2 includes EMMs for main and interaction effects across all outcome analyses.

#### Self-Report Results (SUDS)

The MMANOVA analysis of SUDS used a Toeplitz covariance structure (see the syntax in Supplement) and found a significant main effect of experimental neurostimulation (*F*[2, 373.52] = 19.50, *p* < .000000001). Pairwise comparisons between the three neurostimulation conditions demonstrated that SUDS ratings were significantly lower when HF-rTMS over the dlPFC was administered than when either LF-rTMS over the mPFC (*p <* .00001, *d* = .53) or sham (*p* < .000000001, *d* = .84) were administered. See supplement for additional results for main effects.

A significant interaction was found between experimental neurostimulation, experimental instruction, and group (*F*[22, 705.00] = 18.36, *p* < .000000001). The interaction revealed that participants in the misophonia group experienced significantly more distress than controls when listening (Δ = 1.91, *SE* = .32, *p* < .001) and when downregulating (Δ = 1.93, *SE* = .32, *p* < .001) misophonic sounds across all experimental conditions. In addition, participants in the misophonia group reported significantly less distress than participants with emotion dysregulation when listening to aversive sounds (Δ = 1.76, SE = .32, *p* < .001). In the sham condition alone, participants with emotional dysregulation reported more distress when downregulating aversive sounds than participants with misophonia (Δ = 1.08, SE = .40, *p* < .008). During dlPFC and mPFC stimulation, this difference between groups was no longer significant, suggesting that neurostimulation normalized the regulation of aversive sounds for controls.

Group-specific results were as follows: for participants with emotion dysregulation, HF-rTMS decreased SUDS more than sham when participants were asked to listen to neutral (Δ_SHAM-HF-rTMS_ = 0.92, *p* = 0.004), aversive sounds (Δ_SHAM-HF-rTMS_ = 0.97, *p* < .001, *d* = 0.95), or misophonic sounds (Δ_SHAM-HF-rTMS_ = 1.50, *p* < .001, *d* = 1.26). HF-rTMS also enhanced the downregulation of aversive (Δ_SHAM-HF-tTMS_ = 1.15, *p* < .001, *d* = 0.77) and misophonic (Δ_SHAM-HF-rTMS_ = 1.51, *p* < .001, *d* = 1.28) sounds. LF-rTMS did not outperform sham stimulation in this group (Figure 4). For participants with misophonia, unlike patients with emotion dysregulation, the rTMS effect was specific for the misophonic sounds. There were no differences between both active and sham neurostimulation when participants listened to neutral or aversive sounds or when they downregulated aversive sounds. This is likely because participants with misophonia reported relatively low distress when listening to aversive sounds (*EMM* = 1.55, *SE* = .34). Nevertheless, the distress produced by a misophonic sound was significantly lower in the HF-rTMS condition when compared to sham when participants were instructed to listen (Δ_SHAM-HF-rTMS_ = 0.92, *p* = .001, *d* = 0.74) or to downregulate (Δ_SHAM-HF-rTMS_ = 1.02, *p* < .001, *d* = .94). LF-rTMS was marginally superior to sham during listening (Δ_SHAM-LF-tTMS_ = 0.58, *d* = 0.55, Bonferroni corrected *p* = .16), or downregulation of misophonic sounds (Δ_SHAM-LF-rTMS_ = 0.64, *d* = .59, Bonferroni corrected *p* = .12). There was no difference between the two active rTMS conditions (*p* > .05).

**Figure 4.**
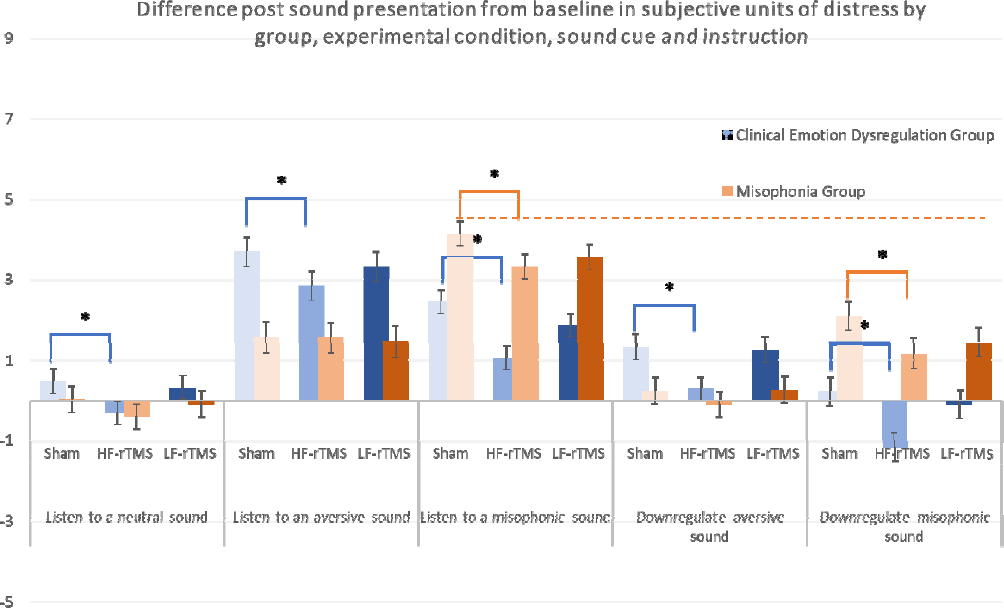
Self-report units of distress by instruction provided, experimental group, and experimental neurostimulation. Values represent estimated marginal means from the mixed models analysis of variance, adjusted for covariates and main effects. The Orange dotted line represents our closest approximation of the misophonic experience.

To examine the effects of implementing a behavioral skill alone, we examined differences within the sham condition only. Instructions to downregulate led to significantly less distress than instructions to listen to aversive (Δ = 1.88, *SE* = .39, *p* < .001) and misophonic (Δ = 2.12, *SE* = .33, *p* < .001) sounds across groups. Thus, in both conditions, excitatory neurostimulation as well as implementation of an emotion regulation skill individually reduced emotional distress, and therefore are promising interventions for both emotional dysregulation and misophonia.

To examine across groups whether neurostimulation alone was superior to behavioral skill implementation, we compared the change in distress when listening to aversive or misophonic sounds while receiving HF-rTMS with changes during sham while participants were engaged in CR. We also compared the combination of neurostimulation with CR with either intervention alone. Utilizing a behavioral skill was superior to neurostimulation alone (*p_aversive_*= .001; *p_misophonic_* = .012) in reducing distress induced by aversive sounds. The combination of neurostimulation and skill use led to significantly lower distress at the end of the sound presentation for both misophonic and aversive sounds when compared to either intervention modality alone (*p*s < .001). Therefore, the combined intervention leads to the highest reduction in distress, followed by utilizing only behavioral skills, followed by neurostimulation alone.

#### Peak arousal (SCR) results

A MMANOVA analysis using a Toeplitz covariance structure found a main effect of baseline (*F*[1, 674.27] = 33.99, *p* < .00000001), the time during the block (*F*[3, 99.85] = 48.86, *p* < .001), and headache (*F*[1, 668.90] = 15.22, *p* < .001). There was also a marginally significant main effect for coil-to-cortex distance (*F*[1, 717.11] = 5.06, *p* = .03). Higher baseline SCR (i.e., during habituation; parameter estimate [PE] = .17, *SE* = .03), lower coil-to-cortex distance (*PE* = −.12, *SE* = .03), and presence of head discomfort (*PE* = −.82, *SE* = .21) led to higher SCR during individual sound presentations. SCR decreased significantly over time within each block (*p*s < .03). A significant main effect of experimental neurostimulation was also found (*F*[2, 587.82] = 6.98, *p* = .001), but pairwise comparisons only found a marginally significant difference between sham and the LF-rTMS conditions (Δ = 0.35, *SE* = .16, *p* = .03). There was no significant main effect for group, instructions provided, or racial background (*p*s > .05). The interaction effect between experimental neurostimulation, instruction provided and experimental group was marginally significant, and as a result, was not investigated further (*F*[22, 1262.55] = 1.65, *p* = .03).

#### Average arousal (SCL) results

A MMANOVA analysis using an unstructured covariance structure found a significant main effect for experimental neurostimulation (*F*[2, 79.70] = 7.18, *p* = .001). Tonic arousal was significantly higher in the sham condition when compared to either active stimulation condition (*p* _HF_rTMS_ = .001; *p*_LF_rTMS_ = .002; *d*s = .14), with no difference between active conditions (*p* > .05) suggesting that both types of neurostimulation reduced tonic arousal. Higher SCL at baseline (i.e., during habituation) led to higher SCL during the experimental runs (*F*[1, 57.77] = 2488.10, *p* < .001). There was also a significant effect of time during the experimental run, with earlier blocks having significantly higher SCL than later blocks (*p*s <.001). Thus, SCL decreased over time during the run independent of the experimental condition. There was no significant main effect for the experimental group, instructions provided, headache, racial background, or coil-to-cortex distance (*p*s > .05).

A significant interaction was found between experimental neurostimulation, instruction provided, and group (*F*[22, 111.50] = 2.69, *p* < .0005, Figure 5). There were no significant differences between groups, and therefore we examined this interaction within each group.

**Figure 5.**
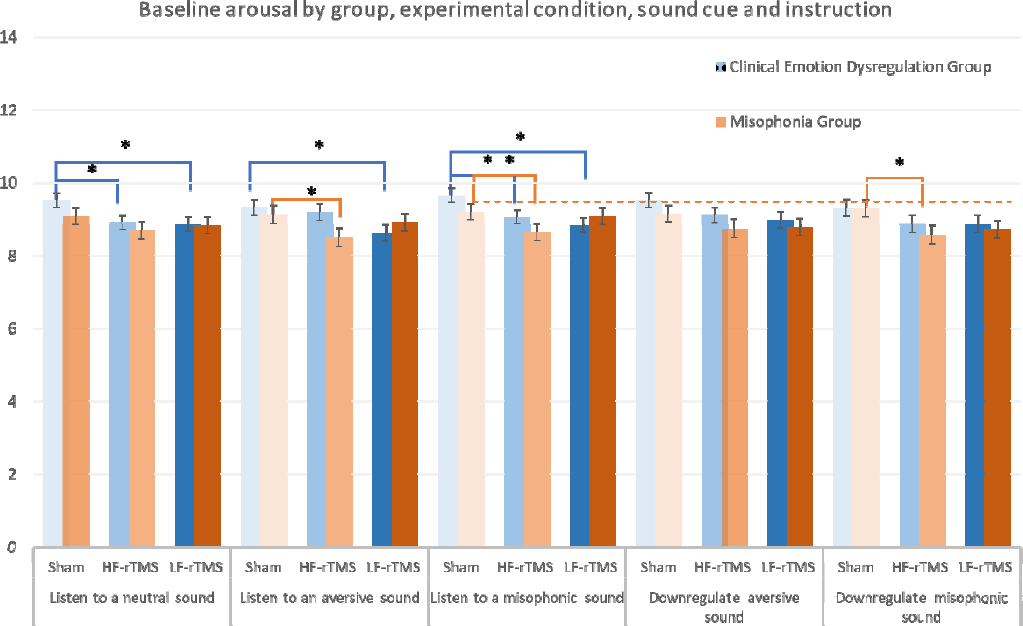
Average skin conductance level in micro siemens depending on experimental group, instruction provided, and experimental neurostimulation. Values represent estimated marginal means from the mixed model analysis of variance, adjusted for covariates and main effects

For clinical control patients with emotion dysregulation, when listening to neutral (Δ_SHAM-HF-rTMS=_ 0.63, *p* = .004; Δ_SHAM-LF-rTMS_ = 0.67, *p* = .002) or misophonic sounds (Δ_SHAM-LF-tTMS_ = 0.811, *p* < .001, *d* = .23; Δ_SHAM-HF-rTMS_ = 0.59, *SE* = 0.203, *p* = .004, *d* = .16), both HF-rTMS and LF-rTMS decreased tonic arousal when compared to sham stimulation, with no difference between the two stimulation conditions. LF-rTMS alone reduced SCL significantly more when listening to an aversive sound when compared to sham (Δ_SHAM-LF-rTMS_ = 0.69, *p* = .002, *d* = .19). There were no significant differences between experimental neurostimulation conditions during the regulation blocks.

For patients with misophonia, HF-rTMS reduced SCL significantly more than sham when listening to either aversive (Δ_SHAM-HF-tTMS_ = 0.63, *p* = .009, *d* = .18) or misophonic sounds (Δ_SHAM-HF-tTMS_ = 0.56, *p* = .009, *d* = .16) and when downregulating misophonic sounds (Δ_SHAM-HF-tTMS_ = 0.73, *p* = .006, *d* = .20). The lowest SCL when exposed to misophonic sounds in the misophonia group was during the combination of behavioral skill utilization and HF-rTMS stimulation.

To investigate the effects of using only behavioral skills, we examined differences within the sham condition and found no significant reduction in SCL when implementing CR alone (i.e., sham stimulation during listening was not significantly different from sham stimulation during downregulation; *p*s >.35). Thus, in both groups, excitatory neurostimulation alone reduced emotional distress.

We also aimed to examine differences across groups in SCL after receiving different intervention approaches while listening to and downregulating aversive and misophonic sounds. Therefore, we compared tonic arousal across three conditions: when participants were downregulating sounds in the sham condition; when participants were listening to sounds and receiving HF-rTMS; when participants were downregulating emotions associated with sounds while receiving HF-rTMS. For both aversive and misophonic sounds, neurostimulation alone (*p_aversive_* = .014; *p_misophonic_*= .014) or in combination with cognitive restructuring (*p_aversive_* = .017; *p_misophonic_* = .001) led to less arousal than CR alone. There was no significant difference between neurostimulation alone and the combined approach (*p*s > .10). There was also no difference between groups in response to these different interventions. This suggests that a neurostimulation-based treatment might be optimal for intervention for misophonia as well as emotional dysregulation.

#### High-Frequency Heart Rate Variability (HF-HRV) Results

A MMANOVA analysis using an unstructured covariance structure did not reveal any main effect of group, instruction provided, headache, or racial background (*p*s > .05). However, we found significant main effects of experimental neurostimulation type (*F*[2, 86.03] = 6.37, *p* = .003). Contrary to our expectations, across both groups and all trials, HF-HRV was significantly higher in the sham condition when compared to either active stimulation condition (*p*_HF_rTMS_ = .008; *p* _LF_rTMS_ = .001, *d*s = .10). In the same analysis, there was a significant main effect of HF-HRV value during habituation (*F*[1, 67.58] = 1088.48, *p* < .00000001) and of coil-to-cortex distance (*F*[1, 121.62] = 11.71, *p* < .0009), with lower coil-to-cortex distance (*PE*= −.02, *SE* = .01) and higher HF-HRV values during habituation (*PE* = .82, *SE* = .03) leading to higher HF-HRV values during experimental trials. Finally, only a marginally significant interaction was found between experimental neurostimulation, instruction, and experimental group (*F*[22, 130.82] = 1.84, Bonferroni corrected *p* = .076), and therefore the interaction was not investigated further.

In sum, when controlling for the increase in HF-HRV induced by neurostimulation or sham alone, participants have higher HF-HRV during the sham experimental trials than during the active stimulation trials. Raw means of HF-HRV across groups are higher for active than for sham neurostimulation (*M_HF-rTMS_* = 0.000186; *M_LF-rTMS_* = 0.000166; *M_sham_* = 0.000153).

### 4. Exit interview

During the exit interview, we asked participants whether the TMS machine produced anxiety, and frustration or made them feel or think differently. There were no differences between groups in how participants experienced neurostimulation (χ = 0.19 - 0.77, *p*s > .1). Across the entire sample, 20-30% experienced emotional distress from the neurostimulation machine, and 33-35% believed the neurostimulation administration altered how they thought and felt. Qualitative data suggested that participants noticed the sound pattern and the tapping from the machine, which contributed to their experience by either increasing their irritability or by providing a different focus for their attention that could distract them from the sounds. In addition, participants perceived response to neurostimulation varied; some commented that neurostimulation either made them more numb to their feelings while others commented that neurostimulation led them to feel things more deeply. Participants with misophonia also described a sense of overwhelming hope, as well as wanting to laugh during the presentation of misophonic triggers because they were much less aversive than when experiencing these stimuli in the absence of neurostimulation.

Participants also reported finding neurostimulation during an emotion regulation task relatively comfortable (*M* = 4.04, *SD* = 2.25, Range: 0 –extremely comfortable – 9 – extremely uncomfortable), with no difference between stimulation conditions (*t*[46.05] = .85, *p* = .40). All participants indicated an above-average willingness to engage in neurostimulation enhanced behavioral treatment to help reduce mental health distress. (*M* = 6.13, *SD* = 2.64, range 0 –unwilling – 9 – extremely willing; *p_group_difference_* = .39). All participants found CR training very helpful (*M* = 7.59, *SD* = 1.83, Range: 0– 9; *p_group_difference_* = .56), the combined procedures highly acceptable *M_acceptability_* = 8.13, *SD* = 1.03, Range 0 – 9; *p_group_difference_* = .68) and reported high likelihood to recommend this treatment to a friend (*M* = 74.98%, *SD* = 19.89, range 0 – 100%; *p_group_difference_* = .88).

## Discussion

Emerging research highlights that misophonia is a serious disorder that significantly impairs the quality of life and functioning [3]. Several studies have characterized misophonia across behavioral and biological domains [5,7,76]; nevertheless, given the intense emotional reaction to specific sounds, it is yet unclear whether misophonia should be considered a unique disorder under the purview of any particular clinical discipline. Furthermore, how to best intervene for those who meet criteria for this disorder continues to be unclear [77]. Therefore, in this study, we compared adults with moderate and severe misophonia severity to a transdiagnostic sample of adults who meet criteria for a DSM-5 disorder and who self-report clinically high difficulties with regulating emotions. We also examined whether interventions should aim to reduce sound reactivity or improve emotion regulation and whether neurostimulation, cognitive restructuring, or their combination offers the most promise to intervene on this disorder.

This study demonstrates that there are few psychophysiological differences in the response and regulation of distress related to sounds between clinical emotional dysregulation and misophonia. Prior preliminary studies did show psychophysiological differences (especially in SCR) between adults with misophonia and non-clinical controls [1]. Our findings highlight that when confronted with trigger sounds, adults with misophonia are similar to adults who have significant difficulties regulating emotions with a range of DSM-5 disorders in physiological reactivity and arousal. Thus, existing evidence-based interventions that successfully address arousal and emotion regulation developed for other clinical problems also may provide some benefit to misophonia sufferers [78].

Significant differences were found in participants’ self-report of distress and regulation. Specifically, clinical controls were more bothered by aversive sounds than those with misophonia; misophonic adults were more upset when compared to controls following triggers and while trying to downregulate emotions induced by trigger sounds. In light of the physiological findings, we interpret this as sensitization: at the perception of a stimulus with which the participant has difficulty, their subjective experience is heightened and doesn’t necessarily mirror the psychophysiological response. An alternative interpretation could be that SCL/SCR/HF-HRV are not appropriate measures to capture the difference in response between groups and other objective measures should be tested to differentiate misophonia from clinical distress. Nevertheless, until a different measure is identified, interventions intended to change misophonic distress should focus on self-reported distress as a unique differentiator from other clinical conditions.

We also examined whether the best way to approach intervention for misophonic distress was to change top-down versus bottom-up processes. Using a functional imaging paradigm, we identified specific functionally connected regions in the anterior insular cortex and medial prefrontal cortex that were more engaged during the presentation of misophonic versus neutral sounds. We examined whether intervening using a bottom-up process, by inhibiting reactivity with the use of neurostimulation, was a potential intervention for misophonia. We also identified the region in the right dorsolateral prefrontal cortex that was most active during regulation when compared to just listening to misophonic sounds. We examined whether a top-down intervention, simulated in the study by enhancing regulation with neurostimulation and/or the use of an emotion regulation skill, could be beneficial for misophonia. Our design allowed us to compare both approaches. We found clear superiority across both self-report and tonic arousal measures for the top-down approach. Participants with misophonia experienced significantly less distress in the presence of misophonic triggers when their regulation network was engaged via either excitatory rTMS over the right dlPFC or the use of an emotion regulation skill. This finding strongly suggests that interventions aimed towards emotion regulation have promise for misophonia.

In the last two decades, neurostimulation has accumulated exciting new evidence that supports its potential for intervention [79]. There are currently five neurostimulation-based protocols that are FDA approved for the treatment of depression, obsessive-compulsive disorder, and smoking [80]. While initially neurostimulation was reserved for treatment-resistant cases, the evidence suggests that non-treatment-resistant adults can significantly benefit from this treatment [81], which opens up this approach to any condition as a candidate novel intervention. In the case of misophonia, no clear pathway for intervention exists yet. Nevertheless, some findings suggest that cognitive behavioral therapy [82,83] and the use of emotion regulation skills [78,84] can offer promise for misophonia. We, therefore, wanted to compare whether neurostimulation alone, emotion regulation alone, or their combination lead to the biggest decrease in distress.

First, it is important to highlight that engaging in either cognitive regulation or neurostimulation alone reduced distress when compared to no intervention for both misophonic and emotionally dysregulated adults. Thus, both neurostimulation and emotion regulation training should be considered important options for future misophonia interventions. Second, the combined intervention on both misophonic and aversive sounds led to significantly less self-reported distress and lower tonic arousal when compared to implementing the emotion regulation skill alone. Self-report results showed the superiority of the behavioral intervention alone versus the neurostimulation alone, while the finding was reversed for the tonic arousal measure. Therefore, if the intended outcome is a reduction in physiological arousal, neurostimulation outperforms behavioral skills. Nevertheless, when it comes to perceived distress, behavioral intervention reduces this perception more so than neurostimulation alone. Given that we found differences primarily in self-reported distress, these findings suggest that, for the optimal misophonia intervention, a combination of neurostimulation and emotion regulation training should be pursued. If a combined treatment is not possible, a behavioral intervention focused on emotion regulation holds more promise than exposing participants to sounds alone.

Our findings also are relevant to the development of emotion regulation interventions in people without misophonia. First, adults with clinical emotional dysregulation had more difficulty in using an emotion regulation skill successfully when compared to misophonic adults. Nevertheless, this difference was no longer significant when neurostimulation was added, suggesting that the addition of TMS can help normalize a dysfunctional process in this clinical sample. In other words, adults with emotion dysregulation may need additional support to implement emotion regulation skills successfully. Furthermore, as with misophonia, the combination of behavioral skills and neuromodulation led to the least distress, suggesting that future interventions should focus on this combined approach for maximum benefit. This finding aligns well with research that highlights the superiority of behavioral therapy in tandem with neurostimulation when compared to behavioral therapy alone [25,85]. The hypothesized mechanism is that neurostimulation facilitates neural processing in localized cortical regions via inducing Hebbian-like plasticity [86] which may have therapeutic effects and may remediate deficits in affected neural networks [87].

Both the behavioral intervention and the neurostimulation approach were feasible to implement and were acceptable to participants. Procedures were well tolerated with some modifications if the initial attempt was uncomfortable, and no participants dropped out of the study once the procedure started. HF rTMS tended to be associated with more headache and scalp discomfort during and immediately after the procedure, although these side effects did not last for more than a day for the majority of participants. Therefore, future neurostimulation trials should consider an analgesic at the site of stimulation (e.g., lidocaine), as well as informing participants that the likelihood of a headache is above average. Our implementation of a sham trial was successful, as participants stayed blinded and did not guess the assigned condition. Beyond acceptability, participants expected the combination to be helpful and promising for a future misophonia intervention. The training in emotion regulation was rated as highly relevant and useful.

There were some key differences between our study and prior studies. First, in our prior examination of psychophysiology during neurostimulation, we found no difference between active and sham rTMS in heart rate variability and indices of arousal [25,51]. In this study, HF-rTMS increased HF-HRV significantly more than sham during the habituation period, and both types of stimulation increased arousal more than sham. This difference may be due to the procedures employed. This study employed an enhanced stimulation regimen with shorter ITT (15 vs. 26 s) and more pulses administered in a shorter amount of time (3 vs 10 min). More intensive excitatory neurostimulation may lead to more effect on HF-HRV, and the shorter duration may not allow sufficient time for behavioral regulation alone to happen. Furthermore, across all experimental conditions, HF-HRV ended up being higher in sham than in active neurostimulation conditions when controlling for the enhanced effect of neurostimulation alone on this measure during habituation. This would suggest that rTMS does not enhance regulation, but rather it facilitates a faster regulation process. Indeed, in our prior trial, where participants regulated for 10 minutes, the sham condition “caught up” and showed similar HF-HRV towards the end of the regulation period, but was significantly different at the beginning of the regulation period [25].

As in other studies [25,73], we found that coil-to-cortex distance was a significant covariate; a larger distance predicted less efficacy of neurostimulation for several outcomes. Greater distance means that the strength of the electric field induced by neurostimulation is reduced by the time it reaches the cortical target, and, therefore, the efficacy of neuromodulation decreases. Furthermore, the presence of headaches increased skin response during experimental trials. Therefore, future research should control for these variables when interpreting outcomes.

These results should be considered in light of several key limitations. While the study was adequately powered, the sample size was small and lacked racial and ethnic diversity across groups. Results should be replicated with a larger sample size and better representation of minoritized populations. LF-rTMS was applied at 90% MT and HF-rTMS was applied at 120% MT per existing protocols at the time when the study was designed. The superiority of HF-rTMS may be due to the higher stimulation intensity. Higher LF-rTMS intensity and alternative ways to target the anterior insula should be investigated before completely discounting the viability of targeting with neurostimulation the anterior insula via its functional connectivity with the mPFC to treat misophonia or emotional dysregulation. Despite our efforts to control for cumulative effects, lingering rTMS effects may have also influenced the results. A between-subjects design would be ideal for a replication study to rule out the influence of cumulative effects, although here we controlled for task order and presented experimental conditions in a randomized order. Lastly, there were several protocol deviations engaged that may confound study results. We included all data available and described our study deviations because the problems we encounter mimic real-world issues and solutions that would need to be implemented in a community practice should an intervention emerge from our protocol.

Taken together, these findings highlight that neurostimulation and cognitive restructuring are promising avenues of intervention for misophonia, with their combination showing the most promise. Engaging in regulation may lead to more reduction in misophonic distress than attempting to inhibit reactivity to sounds. Adults with misophonia have similar psychophysiological responses to stress and triggers as adults with clinical emotional dysregulation, although self-reported distress differentiates groups. This highlights the conclusion that misophonia may be a disorder comparable to other DSM disorders and that additional biomarkers that capture the unique misophonic distress are still needed. It is important to highlight that our study simply probed into different approaches for intervention. Future studies should examine the short and long-term effects of neurostimulation, emotion regulation, or their combination for misophonia.

## Data Availability

All data produced in the present study are available upon reasonable request to the authors

https://doi.org/10.7924/r4ww7jg4k

## Acknowledgments

Data from the present paper were presented as part of several conference talks. This research and the completion of the manuscript were supported by a Misophonia Fund award granted to the first author by the REAM Foundation. The authors would like to thank the participants who took part in this study and acknowledge Lisalynn Kelley, Judith Wright, Brenden Li, Jessica Choi, John Powers, Ph.D., Simon Davis, Ph.D., and our research assistants and the DUMC, BIAC, and BSRC staff for their contributions.

## Data Availability

Data in SPSS and .csv format along with the data dictionary was submitted and accepted on 10/27/2022. It is available currently at the Duke Research Data Repository (RDR): Neacsiu, A., LaBar, K., Rosenthal, M. Z., Bukhari-Parlakturk, N., Kelley, L. (2022). Identifying the optimal neural target for misophonia interventions. Duke Research Data Repository. https://doi.org/10.7924/r4ww7jg4k. Imaging data is available upon request.

## Author Contributions

Conception and design of the study: ADN, KSL, NBP, MZR; acquisition and analysis of data: ADN, LB, NG, VS; drafting the manuscript or figures: LB, NG, MRZ, KSL, NBP.

