## Supplementary material for "Identifying Optimal Parameters for Neuroscience-Informed Interventions for Misophonia": Table 1

**Table 1**. *Demographics and clinical descriptive by group*

|  | Emotion dysregulation (n = 30) | Misophonia  (*n* = 29) | Statistical difference |
| --- | --- | --- | --- |
| Mean age (SD) | 27.07 (8.28) | 29.59 (9.79) | *t*(57) = -1.07, *p* = .29 |
| Female gender identity (%) | 86.67 | 89.66 | *χ^2^*[4] = 2.98, *p* = .56 |
| Latinx background (%) | 23.33 | 3.45 | *χ^2^*[1] = 4.98, *p* = .03 |
| Racial background (%) |  |  | *χ^2^*[5] = 13.68, *p* = .02 |
| Asian/Asian American | 30.0 | 3.45 |  |
| Black/African American | 3.33 | 0.00 |  |
| Native American, American Indian, or Alaskan Native | 3.33 | 0.00 |  |
| White/Caucasian | 56.67 | 96.55 |  |
| Middle Eastern | 6.66 | 0.00 |  |
| On psychotropic medications: | 40.00 |  |  |
| Recent psychotherapy: | 50.00 |  |  |
| Total # of diagnoses, current (SD) | 1.97 (1.38) | 1.52 (1.83) | *t*(57)= 1.07, *p* = .29 |
| Total # of diagnoses, lifetime (SD) | 4.07 (2.12) | 3.00 (2.32) | *t*(57) = 1.85, *p* = .07 |
| Current disorders (%) |  |  |  |
| Mood disorders | 30.00 | 13.80 |  |
| Anxiety disorders | 83.30 | 69.00 |  |
| Obsessive compulsive disorders | 16.70 | 6.90 |  |
| Stress disorders | 10.00 | 10.30 |  |
| Impulse control disorders | 6.70 | 6.90 |  |
| Eating disorders | 0.00 | 3.40 |  |
| Lifetime disorders (%) |  |  |  |
| Mood disorders | 83.30 | 69.00 |  |
| Anxiety disorders | 93.30 | 75.90 |  |
| Substance use disorders | 26.70 | 27.60 |  |
| Obsessive compulsive disorders | 23.30 | 10.30 |  |
| Stress disorders | 31.00 | 20.70 |  |
| Eating disorders | 13.30 | 13.80 |  |
| Any pd (%) | 48.27 | 24.14 | *χ^2^*[1] = 3.66, *p* = .06 |

Note: SD = Standard Deviation; PD = personality disorder
