## Supplementary material for "Identifying Optimal Parameters for Neuroscience-Informed Interventions for Misophonia": Table 2

Table 2: Estimated marginal means (and standard errors) from mixed models analyses by experimental condition and group

A. Main Effects

|  | **∆SUDS** | **SCL** | **SCR** | **HF-HRV** |
| --- | --- | --- | --- | --- |
| **Sham rTMS** | 1.67 (.16) | 9.33 (.15) | 10.03 (.59) | 1.98 (.02) |
| **HF-rTMS** | 0.76 (.17) | 8.84 (.16) | 10.27 (.60) | 1.92 (.02) |
| **Lf -rTMS** | 1.34 (.16) | 8.86 (.15) | 9.68 (.59) | 1.91 (.02) |
| **Listen to neutral** | -0.01 (.19) | 8.99 (.13) | 9.90 (.59) | 1.93 (.02) |
| **Listen to aversive** | 2.43 (.29) | 8.96 (.15) | 10.08 (.61) | 1.92 (.03) |
| **Listen to misophonic** | 2.72 (.19) | 9.09 (.13) | 9.96 (.59) | 1.94 (.02) |
| **Downregulate aversive** | 0.53 (.20) | 9.06 (.14) | 10.04 (.60) | 1.95 (.02) |
| **Downregulate misophonic** | 0.62 (.26) | 8.95 (.15) | 9.97 (.61) | 1.92 (.03) |

B. Interaction effects

|  | | Emotion dysregulation group | | Misophonia group | |
| --- | --- | --- | --- | --- | --- |
|  |  | **∆SUDS** | **SCL** | **∆SUDS** | **SCL** |
| Listen to neutral | **Sham rTMS** | 0.53 (.29) | 9.54 (.20) | 0.06 (.31) | 9.08 (.23) |
|  | **HF rTMS** | -0.39 (.30) | 8.91 (.20) | -0.48 (.32) | 8.70 (.23) |
|  | **LF rTMS** | 0.30 (.30) | 8.87 (.21) | -0.06 (.32) | 8.84 (.23) |
| Listen to aversive | **Sham rTMS** | 3.78 (.37) | 9.33 (.22) | 1.63 (.37) | 9.14 (.25) |
|  | **HF rTMS** | 2.81 (.37) | 9.20 (.22) | 1.50 (.38) | 8.51 (.25) |
|  | **LF rTMS** | 3.33 (.37) | 8.64 (.22) | 1.52 (.31) | 8.92 (.25) |
| Listen to misophonic | **Sham rTMS** | 2.48 (.29) | 9.66 (.20) | 4.17 (.30) | 9.21 (.22) |
|  | **HF rTMS** | 0.98 (.30) | 9.07 (.20) | 3.25 (.32) | 8.65 (.23) |
|  | **LF rTMS** | 1.83 (.29) | 8.85 (.20) | 3.59 (.31) | 9.09 (.22) |
| Downregulate aversive | **Sham rTMS** | 1.36 (.30) | 9.54 (.21) | 0.28 (.32) | 9.16 (.24) |
|  | **HF rTMS** | 0.22 (.31) | 9.12 (.21) | -0.15 (.33) | 8.75 (.25) |
|  | **LF rTMS** | 1.22 (.30) | 8.99 (.22) | 0.28 (.33) | 8.80 (.24) |
| Downregulate misophonic | **Sham rTMS** | 0.28 (.35) | 9.32 (.23) | 2.13 (.34) | 9.31 (.24) |
|  | **HF rTMS** | -1.23 (.35) | 8.87 (.24) | 1.11 (.37) | 8.57 (.27) |
|  | **LF rTMS** | -0.11 (.35) | 8.88 (.24) | 1.50 (.34) | 8.72 (.25) |

*Note.* rTMS = repetitive transcranial magnetic stimulation; HF-rTMS = high frequency rTMS; LF rTMS = low frequency rTMS; SUDS = change in subjective units of distress from baseline; SCL = skin conductance level; SCR = Skin conductance response; HF-HRV = high frequency heart rate variability. EMMs = estimated marginal means computed controlling for baseline and for covariates (coil to cortex distance, headache, racial background)
