## Supplement for "Identifying Optimal Parameters for Neuroscience-Informed Interventions for Misophonia"

***Supplemental Information***

**Methods:**

**Participants and Procedures**

We received online screens from 1341 potential unique participants (79.6% female, 9.2% Hispanic, 79.7% Caucasian), of which 313 adults qualified and could be reached and screened by phone to further examine study inclusion/exclusion criteria. Of these, 177 were invited to an in-person intake assessment. One hundred and twelve unique adults signed consent and participated in an in-person screen to establish diagnostic profile, and to further determine eligibility.

We measured the severity of misophonia and difficulties with emotion regulation on both the online and the in-person screen. Participants were included in the study if their scores fit within pre-determined parameters at the intake assessment. Twenty-two participants were no longer eligible at intake because their scores changed sufficiently to be outside of the pre-determined cutoffs for the misophonic and clinical control groups. In addition, 19 participants were not eligible at the in-person screen (Figure 1), and 12 withdrew, were lost to contact before the experimental day, or couldn’t do the MRI visit because of claustrophobia or technical difficulties. Thus, we enrolled 59 participants into the study (enrolled group) of which 54 participants (completer group) were present for the neurostimulation experimental day (27 in each group).

Enrolled participants were 4 men, 52 women, and 3 non-binary adults between the ages of 18 and 54 (*M* = 28.31; *SD* = 9.06 years) who self-reported significant misophonic severity, or who met criteria for any DSM-5 disorder (excluding active substance use, psychotic disorders, and Bipolar I) and reported above-average emotional dysregulation. Participants met the criteria for an average of 1.75 (*SD =* 1.62) current diagnoses and 3.54 (*SD* = 2.26) lifetime diagnoses according to the structured interview for *DSM-5* disorders (SCID-5) [1] (Table 1). Twenty-one participants (36.21%) met criteria for at least one personality disorder according to the SCID-5-PD [2].

**Measures**

*Diagnostic Assessment:* The SCID-5 [1] and SCID-PD [2] have demonstrated high diagnostic accuracy (83%) and strong inter-rater reliability (.85 during training and .76 at a Quality Assurance check) [3]. Participants were led through both structured interviews by either the first author (65.5% of cases) or one of two trained diagnostic assessors under the supervision of the first author. In the cases where the first author did not conduct the interview, she reviewed in detail with the assessor the questions asked to confirm the diagnostic profile. In case of disagreement, she reassessed the disorder at the next visit.

*Difficulties in Emotion Regulation Scale* (DERS): The DERS [4] is a 36-item instrument that assesses typical levels of emotion dysregulation across six domains (awareness, nonacceptance, strategies, goal-oriented behaviors, impulsivity, and emotional clarity). Participants respond on a Likert scale ranging from 1 (almost never) to 5 (almost always). In the original measure development paper, the DERS was found to have high internal consistency (α = .93), good test-retest reliability (r = .88, p < .01), and adequate construct and predictive validity. The total score is the sum of all items (some reversed). Higher scores indicate more dysregulation. In the present study, Cronbach’s alpha at intake was .84, indicating acceptable internal consistency.

*Misophonia Questionnaire (MQ).* The MQ is a three-part assessment of the types of misophonic sounds the participant is sensitive to and the symptoms experienced. Part one is an 8-item scale that assesses the participant’s sensitivity in comparison to others of seven common misophonic triggers and one item for “other.” The items use a 5-point Likert scale to identify to what degree the participant is sensitive to each, ranging from 0 (not at all true) to 4 (always true). Part two is completed only if the participant scores a 1 (rarely true) on any of the items in part one. Part two is an 11-item scale that assesses the frequency of ten reactions and one “other” reaction by the participant once they are aware of the misophonic sounds from part one. The items use a 5-point Likert scale to identify the frequency of each reaction, ranging from 0 (never) to 4 (always). Part three is a 15-point self-rating scale of sound sensitivity. There are 5 items with 3 points associated with each with responses ranging from 1-3 (minimal within range of normal or very mild sound sensitivities) to 13-15 (very severe sound sensitivities). Previous studies using the MQ have reported strong reliability and evidence of construct validity [5]. In the present study, Cronbach’s alpha was .85 for part I and .92 for part II, indicating acceptable internal consistency.

*Positive and Negative Affect Scale* (PANAS). The PANAS [6] is a 28-item assessment of the degree to which the participant has felt a wide range of positive and negative emotions and feelings on the day of the assessment. The items use a 5-point Likert scale ranging from 1 “Not at all or very slightly” to 5 “Extremely” concerning the extent the participant has experienced each emotion. Cronbach’s alpha before the intake session was .88.

*Exit Interview:* Questions about feasibility included difficulty with limiting movement, level of comfort, ability to concentrate given the TMS noise, distress about the procedures, ease to hear and understand the clinician, connection with the clinician, and session engagement. Topics in questions about acceptability included session length, skills training, TMS procedures, personalized stressors use, and ambulatory phone assessment. Feasibility and acceptability questions were rated on a scale from 0 (not at all) to 9 (extremely) and scores were reversed as needed and averaged to compute an overall acceptability score where 0 represented not feasible/acceptable at all and 9 represented very feasible/acceptable. Satisfaction was rated on a 0 (low) to 100 (high) continuous scale.

**Sound Task**

The task was programmed in MATLAB. Neutral sounds were randomly selected from all sounds that were marked as neutral. If there were too many high-arousal sounds, a randomizer was employed to select the stimuli. If there were too few high-arousal sounds, a randomizer was employed to select the sound(s) that repeated once within the stimulus set to achieve a total of 12 stimuli. More stimuli were not presented in this case because of participant fatigue (participants had already listened to over 100 possible stimuli).

One participant completed the task twice due to technical difficulties recording sounds the first time. Another participant came too late to complete their sound task, which could not be rescheduled, and as a result, another participant’s personalized stimulus set (from the same group) was used to run them through the experimental sessions. Post-hoc examination of their data did not indicate any deviations from the group data, and therefore their results were included in the analyses.

**Neuroimaging Session**

***MRI acquisition:***

Participants viewed the screen via a mirror system located on the head coil and the start of each run was electronically synchronized with the MRI acquisition computer. Behavioral responses were recorded with a 4-key fiber-optic response box (Resonance Technology, Inc.). When necessary, vision problems were corrected using MRI-compatible lenses that matched the distance prescription used by the participant.

#### Neuroimaging Data Analysis:

Preprocessing of the high-resolution, T1-weighted anatomical images included intensity correction using N4BiasFieldCorrection [7], skull-stripping, and spatial normalization to the ICBM 152 Nonlinear Asymmetrical template v2009c [8]. Initial preprocessing of the functional image data included slice time correction, calculation of motion correction transforms, and calculation of the transform for spatial registration to the high-resolution T1-weighted image. The fMRI data were then moved to standard space by applying the concatenated motion-correction transforms, fMRI-to-T1 transform, and T1-to-ICBM transform.

Model regressors were created for the fixation cross (3-7s), active baseline arrow task (9s), cued “listen” instruction (5s), cued “downregulate” instruction (5s), “listen” strategy during a neutral sound (“Hear Neutral”, 10s), “listen” strategy during an aversive sound (“Hear Aversive”, 10s), “listen” strategy during a misophonic sound (“Hear misophonic”, 10s), “downregulate” strategy during an aversive sound (“Downregulate Aversive”; 10s), “downregulate” strategy during a misophonic sound (“Downregulate Misophonia”; 10s), and distress ratings (10 s). A weight of 1 was attributed to each task regressor in the general linear model (GLM). The trial “on” times were convolved with a double-gamma hemodynamic response function to create the final GLM regressors.

A generalized psychophysiological interaction analysis was conducted to measure left insula connectivity with the left mPFC. The time course of the left insula as the seed was the physiological variable, and the BOLD activation during the “listen to misophonic vs. neutral sound” contrast was used as the psychological regressor.

### Neurostimulation Experimental Session

***Skills Training:***

In teaching *distancing*, adopting a detached and unemotional attitude was introduced as objective distancing [9]. Distancing by using time and space was also discussed and practiced on standardized examples [10]. For the misophonia group, distancing was further refined as thinking about what else could be making that sound (to have a different image in mind), thinking that the sound is time-limited, or thinking that, in the context of one’s day, the misophonic experience is but a small percentage of the time. We also discussed space distancing, trying to focus one’s attention on another element of the situation that is not as upsetting (i.e., not the visual stimulus that produces the trigger sound).

Participants were also taught *reframing* using an adapted version of existing paradigms [11-13].

Specifically, we emphasized with pictures and examples the relationship between thoughts and emotions in ambiguous situations and identified effectiveness as the objective in an emotional situation as opposed to being right. Participants were instructed to find less toxic interpretations to be effective rather than right when upset. Participants learned to think about elements of the situation that they did not pay attention to or information that was missing and to reframe their cognitions based on the full picture with an eye toward effectiveness. Participants were also taught to examine the worst-case scenario, the probability of it occurring, and the likelihood of survival if it occurred.

For the misophonia group, we worked on reframing thoughts related to the person producing the sound (“they’re hungry rather than they’re rude”; “they’re anxious rather than they’re annoying me on purpose”). We also discussed using the three questions (worst-case scenario, likelihood it will happen, and likelihood of survival) during an experience where misophonic triggers are present. The CR training ended with a quiz that included definitions as well as fake scenarios to practice for the clinician to ensure that the skills were understood and could be applied appropriately.

#### TMS experiment

To measure psychophysiological measurements continuously during the experiment, GSR Ag/AgCl electrodes filled with an isotonic gel were placed on the distal phalanges of the index and ring fingers. HR electrodes were placed on the ankle and wrist. Amplified analog data were converted to digital recording and filtered using BIOPAC’s AcqKnowledge 4.1 software.

For both the right dlPFC and the right mPFC targets, 87% of z-scores were above a significance threshold of z = 1.96. DlPFC targets were on average 14.23 mm away from the skull (SD = 1.89mm, range 9.8 mm – 18.4 mm), similar to mPFC targets (M = 14.94 mm, SD = 1.93, range = 10.4 – 18.8 mm). Stimulation intensity ranged from 38 to 97% from maximum stimulator output (MSO) for the dlPFC target (M = 56.19, SD = 10.75), and from 29 to 73% MSO for the mPFC target (M = 42.19, SD = 8.10).

***Statistical Analyses***

The syntax for analyses on primary outcomes during the habituation period alone:

MIXED lg_Habit_HF_HRV BY targetRegion run group headache_bin race_bin WITH coilToBrain lg_taskBaseline_HF_HRV

/CRITERIA = CIN(95) MXITER(100) MXSTEP(5) SCORING(1)

SINGULAR(0.000000000001) HCONVERGE(0, ABSOLUTE) LCONVERGE(0, ABSOLUTE)

PCONVERGE(0.000001, ABSOLUTE)

/FIXED = targetRegion run group headache_bin coilToBrain race_bin lg_taskBaseline_HF_HRV group*targetRegion | SSTYPE(3)

/METHOD = REML

/PRINT = SOLUTION TESTCOV R

/REPEATED = run block | SUBJECT(SubjectID) COVCONDITION(un)

/EMMEANS = TABLES(targetRegion) COMPARE ADJ(LSD)

/EMMEANS = TABLES(race_bin) COMPARE ADJ(LSD)

/EMMEANS = TABLES (Group) COMPARE ADJ(LSD)

/EMMEANS = TABLES (group*targetRegion) COMPARE (group)

/EMMEANS = TABLES (group*targetRegion) COMPARE (targetRegion).

MIXED habit_maxGSR BY targetRegion run group headache_bin race_bin WITH coilToBrain taskBaseline_maxGSR

/CRITERIA = CIN(95) MXITER(100) MXSTEP(5) SCORING(1)

SINGULAR(0.000000000001) HCONVERGE(0, ABSOLUTE) LCONVERGE(0, ABSOLUTE)

PCONVERGE(0.000001, ABSOLUTE)

/FIXED = targetRegion run group headache_bin coilToBrain race_bin taskBaseline_maxGSR group*targetRegion | SSTYPE(3)

/METHOD = REML

/PRINT = SOLUTION TESTCOV R

/REPEATED = run block | SUBJECT(SubjectID) COVCONDITION(un)

/EMMEANS = TABLES(targetRegion) COMPARE ADJ(LSD)

/EMMEANS = TABLES (Group) COMPARE ADJ(LSD)

/EMMEANS = TABLES (group*targetRegion) COMPARE (group)

/EMMEANS = TABLES (group*targetRegion) COMPARE (targetRegion).

MIXED habit_avgSCL BY targetRegion run group headache_bin race_bin WITH coilToBrain taskBaseline_avgSCL

/CRITERIA = CIN(95) MXITER(100) MXSTEP(5) SCORING(1)

SINGULAR(0.000000000001) HCONVERGE(0, ABSOLUTE) LCONVERGE(0, ABSOLUTE)

PCONVERGE(0.000001, ABSOLUTE)

/FIXED = targetRegion run group headache_bin coilToBrain race_bin taskBaseline_avgSCL group*targetRegion | SSTYPE(3)

/METHOD = REML

/PRINT = SOLUTION TESTCOV R

/REPEATED = run block | SUBJECT(SubjectID) COVCONDITION(un)

/EMMEANS = TABLES(targetRegion) COMPARE ADJ(LSD)

/EMMEANS = TABLES (Group) COMPARE ADJ(LSD)

/EMMEANS = TABLES (group*targetRegion) COMPARE (group)

/EMMEANS = TABLES (group*targetRegion) COMPARE (targetRegion).

MIXED

delta_suds BY targetRegion run instruction group block race_bin timepoint headache_bin WITH

coilToBrain

/CRITERIA = CIN(95) MXITER(100) MXSTEP(5) SCORING(1)

SINGULAR(0.000000000001) HCONVERGE(0, ABSOLUTE) LCONVERGE(0, ABSOLUTE)

PCONVERGE(0.000001, ABSOLUTE)

/FIXED = targetRegion run instruction block group timepoint race_bin headache_bin coilToBrain group*targetRegion*instruction | SSTYPE(3)

/METHOD = REML

/PRINT = SOLUTION TESTCOV R

/REPEATED = run block timepoint | SUBJECT(SubjectID) COVCONDITION(tp)

/EMMEANS = TABLES( instruction) COMPARE ADJ(LSD)

/EMMEANS = TABLES (Group) COMPARE ADJ(LSD)

/EMMEANS = TABLES (block) COMPARE ADJ(LSD)

/EMMEANS = TABLES (group*targetRegion*instruction) COMPARE (group)

/EMMEANS = TABLES (group*targetRegion*instruction) COMPARE (instruction).

Syntax for primary outcome analyses (using habituation values instead of baseline values given significant differences during habituation):

MIXED delta_suds BY targetRegion instruction run group block race_bin timepoint headache_bin WITH

coilToBrain

/CRITERIA = CIN(95) MXITER(100) MXSTEP(5) SCORING(1)

SINGULAR(0.000000000001) HCONVERGE(0, ABSOLUTE) LCONVERGE(0, ABSOLUTE)

PCONVERGE(0.000001, ABSOLUTE)

/FIXED = targetRegion run instruction block group timepoint race_bin headache_bin coilToBrain group*targetRegion*instruction | SSTYPE(3)

/METHOD = REML

/PRINT = SOLUTION TESTCOV R

/REPEATED = run block timepoint | SUBJECT(SubjectID) COVCONDITION(tp)

/EMMEANS = TABLES (group*targetRegion*instruction) COMPARE (instruction)

/EMMEANS = TABLES (group*targetRegion*instruction) COMPARE (targetRegion)

/EMMEANS = TABLES (group*targetRegion*instruction) COMPARE (Group)

/EMMEANS = TABLES (targetRegion*instruction) COMPARE (instruction)

/EMMEANS = TABLES (group*instruction) COMPARE (group).

MIXED maxGSR BY targetRegion run instruction group block race_bin timepoint headache_bin WITH

coilToBrain habit_maxGSR

/CRITERIA = CIN(95) MXITER(100) MXSTEP(5) SCORING(1)

SINGULAR(0.000000000001) HCONVERGE(0, ABSOLUTE) LCONVERGE(0, ABSOLUTE)

PCONVERGE(0.000001, ABSOLUTE)

/FIXED = targetRegion run instruction block group timepoint race_bin headache_bin habit_maxGSR

coilToBrain group*targetRegion*instruction | SSTYPE(3)

/METHOD = REML

/PRINT = SOLUTION TESTCOV R

/REPEATED = run block timepoint | SUBJECT(SubjectID) COVCONDITION(tp)

/EMMEANS = TABLES(targetRegion) COMPARE ADJ(LSD)

/EMMEANS = TABLES(instruction) COMPARE ADJ(LSD)

/EMMEANS= TABLES (Group) COMPARE ADJ(LSD)

/EMMEANS= TABLES (timepoint) COMPARE ADJ(LSD)

/EMMEANS = TABLES (group*targetRegion*instruction) COMPARE (instruction)

/EMMEANS = TABLES (group*targetRegion*instruction) COMPARE (targetRegion)

/EMMEANS = TABLES (group*targetRegion*instruction) COMPARE (Group)

/EMMEANS = TABLES (targetRegion*instruction) COMPARE (instruction)

/EMMEANS = TABLES (group*instruction) COMPARE (group).

MIXED lg_HF_HRV BY targetRegion instruction run group block headache_bin race_bin WITH coilToBrain lg_Habit_HF_HRV

/CRITERIA = CIN(95) MXITER(100) MXSTEP(5) SCORING(1)

SINGULAR(0.000000000001) HCONVERGE(0, ABSOLUTE) LCONVERGE(0, ABSOLUTE)

PCONVERGE(0.000001, ABSOLUTE)

/FIXED = targetRegion instruction run block Group lg_Habit_HF_HRV headache_bin race_bin coilToBrain group*targetRegion*instruction | SSTYPE(3)

/METHOD = REML

/PRINT = SOLUTION TESTCOV R

/REPEATED = run block | SUBJECT(SubjectID) COVCONDITION(un)

/EMMEANS = TABLES(targetRegion) COMPARE ADJ(LSD)

/EMMEANS = TABLES(instruction) COMPARE ADJ(LSD)

/EMMEANS = TABLES (Group) COMPARE ADJ(LSD)

/EMMEANS = TABLES (block) COMPARE ADJ(LSD)

/EMMEANS = TABLES (group*targetRegion*instruction) COMPARE (instruction)

/EMMEANS = TABLES (group*targetRegion*instruction) COMPARE (targetRegion)

/EMMEANS = TABLES (group*targetRegion*instruction) COMPARE (Group)

/EMMEANS = TABLES (targetRegion*instruction) COMPARE (instruction)

/EMMEANS = TABLES (group*instruction) COMPARE (group).

MIXED avgSCL BY targetRegion instruction run group block headache_bin race_bin WITH coilToBrain habit_avgSCL

/CRITERIA = CIN(95) MXITER(100) MXSTEP(5) SCORING(1)

SINGULAR(0.000000000001) HCONVERGE(0, ABSOLUTE) LCONVERGE(0, ABSOLUTE)

PCONVERGE(0.000001, ABSOLUTE)

/FIXED = targetRegion instruction run block group habit_avgSCL headache_bin race_bin coilToBrain group*targetRegion*instruction | SSTYPE(3)

/METHOD = REML

/PRINT = SOLUTION TESTCOV R

/REPEATED = run block | SUBJECT(SubjectID) COVCONDITION(un)

/EMMEANS = TABLES(targetRegion) COMPARE ADJ(LSD)

/EMMEANS = TABLES(instruction) COMPARE ADJ(LSD)

/EMMEANS= TABLES (Group) COMPARE ADJ(LSD)

/EMMEANS= TABLES (block) COMPARE ADJ(LSD)

/EMMEANS = TABLES (group*targetRegion*instruction) COMPARE (instruction)

/EMMEANS = TABLES (group*targetRegion*instruction) COMPARE (targetRegion)

/EMMEANS = TABLES (group*targetRegion*instruction) COMPARE (Group)

/EMMEANS = TABLES (targetRegion*instruction) COMPARE (instruction)

/EMMEANS = TABLES (group*instruction) COMPARE (group).

**Results:**

**Feasibility, acceptability, tolerability, compliance, and blinding efficacy**

Acceptability and feasibility: Once the MRI or the neurostimulation experimental session was started, none of the participants withdrew from the study, supporting the self-reported high levels of acceptability for the proposed procedures. Overall participants in both groups reported reduced stress at the end compared to the beginning of each experimental session (∆_average_intake_change_ = -0.56, SD = 1.53; ∆_average_MRI_change_ = -0.31, SD = 1.22; ∆_average_TMS_change_ = -0.48, SD = 0.89). There were no differences in stress reduction over time or between the experimental group (*F_time_* [2] = 0.09, *p* = .92; *F_group_* [1] = 0.19, *p* = .67). The PANAS showed no significant change in negative affect from before to after the sound task (*F_time_* [1] = 1.03, *p* = .31). Participants in the misophonia group reported more negative affect than controls during the sound (*F_group_* [1] = 5.39, *p* = .02) and MRI (*F_group_* [1] = 4.93, *p* = .03) tasks. There was a significant decrease over time in negative affect following the MRI (*F_time_* [1] = 4.65, *p* = .04) and the neurostimulation experiments (*F_time_* [1] = 15.65, *p* < .001). A significant interaction effect showed that only participants in the misophonia group dropped in their negative affect over time significantly (∆_change_MRI_ = 1.93, SE = 0.61, *p* = .003). Overall, these results suggest that the experimental tasks were feasible and acceptable to our participants.

Tolerability: There were no significant differences in neck pain between trials and between groups (*p*s > .36), with 52.83% of trials resulting in some neck pain. More participants in the HF-rTMS trial (44.44%) than in the LF-rTMS (33.33%) and sham (18.52%) trials experienced some scalp discomfort at the end of the trial (*χ^2^*[2] = 8.38, *p* = .015). For those who experienced scalp discomfort, the severity on a scale from 0 (not uncomfortable at all) to 9 (extremely painful) was mild (*M* = 2.35, *SD* = 1.67). Similarly, the severity of headache (*M* = 2.64, *SD* = 1.76) and neck pain (*M* = 2.30, *SD* = 1.45) was mild. Scalp discomfort was reported by the same participants who reported headaches, and therefore, it was not added as a data-driven covariate in analyses.

For most of the participants, side effects improved by the end of the experimental session. At the end of the experimental day, seven participants (13%, all in the misophonia condition, *χ^2^*[1] = 8.04, *p* = .005) continued to report a worsening headache when compared to before the experimental session. Only one participant reported worsening neck pain from the beginning of the session (1.9%) and one participant reported neck pain improvement following the session. Four participants (7.4%) continued reporting scalp discomfort by the end of the intervention day. There were no differences between conditions in neck and scalp discomfort (*p*s >.05). There were no reported hearing impairments across groups before or after the session. All participants who left the experimental session with some of these expected side effects reported that they resolved within the next 24 hours and did not have any long-lasting consequences.

Compliance: After each experimental task (sounds, MRI, neurostimulation), we asked participants whether they were distracted or present and how successful they were in following the instructions. Participants reported staying present during the experimental tasks (*M* = 7.18, *SD* = 1.75, range 0-9), and being successful in following the instructions (*M* = 7.50, *SD* = 1.60, range: 0-9) with no differences between groups, or between the type of neurostimulation received (active HF-rTMS, active LF-rTMS, or sham), ps > .17.

Blinding efficacy: Participants were asked after each run whether they believed they received active or sham neurostimulation. Participants were overall unsure, with a tendency to believe they received active stimulation across runs (*M* = 6.30, *SD* = 2.21, Range: 0- I received sham; 5-unsure; 9 – I received active rTMS). There was no difference in guessing active versus sham assignments between types of neurostimulation received (*M_sham_* = 5.72, *SD* = 2.27; *M_HF-rTMS_*= 6.74, *SD* = 2.07; *M_LF-rTMS_* = 6.43, *SD* = 2.18) or between groups (F [2, 151] = 11.05, *p* = .09), according to a multivariate ANOVA controlling for racial background (F [1, 151] = 0.06, *p* = .44).

**Missing data and deviations from the original protocol:**

Of the 59 participants who were enrolled, 54 completed the neurostimulation experiment. Three participants (one with misophonia who received sham, one with misophonia who received HF-rTMS, and one clinical control who received LF-rTMS) found one of the neurostimulation conditions too uncomfortable to complete. Therefore, 96.30% of participants with misophonia completed HF-rTMS and sham, and 100% completed LF-rTMS. In the control group, 100% completed HF-rTMS and sham and 96.30% completed LF-rTMS.

Several protocol deviations involving targeting, stimulation intensity, and ratings occurred to accommodate obstacles with participation (see supplemental file).

**Targeting deviations**: Three participants in the control group did not show any significant right dlPFC activation when contrasting downregulating with hearing misophonic sounds. As a result, the “downregulating vs. hearing aversive sounds” contrast was successfully used instead; these runs were included in analyses. One participant’s mPFC target was located in a suboptimal region due to an analysis error (z score at the mPFC target after corrected analysis was = -.63), and one participant received direct versus reverse current over their mPFC target because of an administrative error (which may have reduced the potency of neurostimulation for this participant for this condition). These two runs were excluded from analyses. For one participant, no right mPFC target could be found; the PI decided to use a left mPFC target and continue with the same protocol. For one participant the right mPFC target was uncomfortable, and as a result, an alternative left mPFC target was used for the LF-rTMS condition. These two runs with left-side targets were also included in analyses.

**Stimulation intensity deviations**: Neurostimulation started below the desired stimulation intensity for 34 out of the 54 participants during the habituation period but was successfully ramped up for the majority of participants by the first experimental block. Thirteen participants (24%) started at least one of their experimental blocks below the desired intensity, and 18.52% completed an entire experimental run below the identified target. For those who did not receive neurostimulation at the desired intensity, rTMS was administered on average at 88.05% of the optimal desired intensity (SD = 7.51, Range: 65.91-97.98). The reasons to stay below desired intensity were related to discomfort during HF-rTMS (7 participants), LF-rTMS or sham (one participant each), or to an excessively high MT leading to the machine overheating (1 participant). Two participants found at least one of the three neurostimulation conditions too painful and did not complete that condition, although they did complete the other two conditions (one had stimulation below target in one run). All available data were included in analyses, including runs where stimulation was below optimal intensity.

**Deviations in self-report measures**: Two clinical control participants rated their SUDS as ‘0’ for all of the sound trials, baselines, and postline (69 ratings). Their data were excluded from the SUDS analysis for either being incorrect or an outlier.

During the neurostimulation experiment, five participants (3 from the emotion dysregulation group and 2 from the Misophonia group) did not indicate their SUDS at the end of one of the task baselines (3.13% of the data points). In these cases, SUDS after the session baseline were used in analyses. Seven participants missed entering SUDS for one of the sounds presented (0.22% of data points), and, as a result, these time points could not be included in the SUDS analysis.

**Expectancies of improvement**

All participants believed combining neurostimulation with CR is likely to improve emotional dysregulation (*M* = 6.74, *SD* = 1.36, range 1-9) without any differences between groups (*t*[52] = 0.20, *p* = .84), and expressed-above average confidence that the behavioral training would improve their difficulties managing emotional distress (*M* = 6.67, *SD* = 1.49, range 1-9). Participants in the emotion regulation control group were marginally more confident in their ability to use CR for emotional distress than their misophonic counterparts (*t*[52] = 2.07, *p* = .04).

Overall, across participants and at the end of the experimental session, expectancies for emotion regulation change were above-average for a future intervention using the tools employed in the study (*M* = 6.64, *SD* = 1.25, range 1-9), without any differences between stimulation conditions (*t*[52] = 1.24, *p* = .22). Participants in the misophonia group also reported above-average expectations that the combination of neurostimulation and cognitive restructuring will reduce misophonic distress (*M* = 6.52, *SD* = 1.90, range 1-9), and expressed above-average confidence that they could use the behavioral skill to reduce misophonic distress (*M* = 6.68, *SD* = 1.96, range 1-9). Overall expectations for the promise of a combined intervention for misophonia were also above average (*M* = 6.59, *SD* = 1.17, range 1-9).

After each experimental task, we asked participants how much they believed neurostimulation helped them with their emotional management. There were no perceived differences between types of neurostimulation, or between groups in the regulation utility of this tool (*p*s > .23), with participants on average rating the helpfulness of neurostimulation as being above-average (*M* = 5.70, *SD* = 2.15, range = 0 – not helpful – 9 – extremely helpful).

As part of the exit interview, we asked participants to report which was the most helpful CR instruction for them. Participants in both groups found both distancing and reframing techniques helpful. For participants with misophonia specifically, the most frequently mentioned preferred strategies were: (1) to tell oneself that the sound/negative experience is temporary and will not last forever; and (2) to mentally change the context of the sound by thinking about what else could make it, characterizing the pitch and tone is and considering other sounds that would have the same pitch and tone, and breaking the sound down in bits that are not connected.

#### Self-Report Additional Results (SUDS)

LF-rTMS over the mPFC also led to a marginally significant reduction in SUDS when compared to sham (Bonferroni corrected *p* = .072, *d* = .31).

A significant main effect of experimental instruction was also found (*F*[4, 490.81] = 106.61, *p* < .000000001), showing that listening to aversive sounds (*p <* .00000001, *d* = 2.24) or misophonic sounds (*p <* .00000001, *d* = 2.51) resulted in significantly higher distress than listening to neutral sounds. There was no difference in distress between listening to misophonic versus aversive sounds across participants (*p* > .05). Downregulation instructions promoted a significant decrease in SUDS compared to simply listening to either aversive (∆_AVERSIVE_ = 1.90, *SE* = 0.37, *p* < .0000001, *d* = 1.75) or misophonic sounds (∆_MISOPHONIC_ = 2.10, *SE* = 0.31, *p* < .000000001, *d* = 1.93). There was no significant difference in distress between the downregulation of misophonic or aversive sounds, suggesting that participants were able to downregulate their emotions following the instruction provided. Finally, a main effect of time during each instruction block (*F*[3, 106.79] = 4.02, *p* = .009) was found. Specifically, the last sound in the block elicited more distress than the first sound in the block (∆ = .22, *SE* = .07, *p* < .001). There was no significant difference between the experimental group, run order, or any of the covariates included (*p*s > .05).
